# Translational Evidence for Dopaminergic Alteration of Basal Ganglia Functional Connectivity in Persons with Schizophrenia

**DOI:** 10.1101/2025.03.31.25324962

**Authors:** Philip N. Tubiolo, John C. Williams, Roberto B. Gil, Clifford Cassidy, Natalka K. Haubold, Yash Patel, Sameera K. Abeykoon, Zu Jie Zheng, Dathy T. Pham, Najate Ojeil, Kelly Bobchin, Eilon B. Silver-Frankel, Greg Perlman, Jodi J. Weinstein, Christoph Kellendonk, Guillermo Horga, Mark Slifstein, Anissa Abi-Dargham, Jared X. Van Snellenberg

**Affiliations:** Department of Biomedical Engineering, Stony Brook University, Stony Brook, NY 11794; Department of Psychiatry and Behavioral Health, Renaissance School of Medicine at Stony Brook University, Stony Brook, NY 11794; Scholars in BioMedical Sciences Training Program, Renaissance School of Medicine at Stony Brook University, Stony Brook, NY 11794; Medical Scientist Training Program, Renaissance School of Medicine at Stony Brook University, Stony Brook, NY 11794; Department of Psychiatry, Columbia University Vagelos College of Physicians and Surgeons, New York-Presbyterian / Columbia University Irving Medical Center, New York, NY 10032; New York State Psychiatric Institute, New York, NY 10032; College of Medicine, State University of New York Downstate Health Sciences University, Brooklyn, NY 11203; Department of Neurobiology and Behavior, Cornell University, Ithaca, NY 14853; Department of Molecular Pharmacology & Therapeutics, Vagelos College of Physicians and Surgeons, New York-Presbyterian / Columbia University Irving Medical Center, New York, NY 10032; Department of Radiology, Renaissance School of Medicine at Stony Brook University, Stony Brook, NY 11794; Department of Psychology, Stony Brook University, Stony Brook, NY 11794

## Abstract

In prior work, a transgenic mouse model of striatal dopamine dysfunction observed in persons with schizophrenia (PSZ) exhibited dopamine-related neuroplasticity of axonal projections in basal ganglia, a phenotype that has never been demonstrated in human PSZ. Consequently, we sought to identify a dopamine-related alteration of basal ganglia connectivity via working memory task-based and resting-state functional magnetic resonance imaging (fMRI), neuromelanin-sensitive MRI (NM-MRI), and positron emission tomography (PET), in unmedicated PSZ. In this case-control study, 37 unmedicated PSZ and 30 demographically matched healthy controls (HC) underwent resting-state fMRI; a subset of 29 PSZ and 29 HC also underwent working memory task-based fMRI, and another subset of 22 PSZ and 20 HC underwent NM-MRI. Primary outcome measures included: 1) task-state and resting-state functional connectivity (FC) between dorsal caudate (DCa) and globus pallidus externus (GPe), and 2) NM-MRI contrast-to-noise ratio in substantia nigra/ventral tegmental area voxels associated with psychotic symptom severity. PSZ displayed elevated DCa-GPe task-state FC, which was associated with increased NM-MRI contrast-to-noise ratio in the substantia nigra and worse working memory task performance. This in-vivo evidence of a dopamine-associated neural abnormality of DCa and GPe FC in unmedicated PSZ suggests a potential neurodevelopmental mechanism of working memory deficits in schizophrenia, which could be a critical step towards developing treatments for cognitive deficits.

## 1. Introduction

Evidence of striatal dopamine dysfunction in persons with schizophrenia (PSZ) is among the most robust and consistently replicated findings in the neurobiology of schizophrenia. Studies with [^18^F]DOPA show elevated presynaptic striatal dopamine synthesis in medication-naïve and medication-free PSZ^1, 2^, as well as individuals at clinical high risk for schizophrenia^3, 4^. Studies using radiolabeled dopamine D2 receptor (D2R) antagonists [^123^I]IBZM or [^11^C]raclopride show increased amphetamine-induced tracer displacement in striatum in PSZ^5, 6^, while depletion of dopamine with alpha-methyl-para-tyrosine results in increased tracer binding in PSZ^7, 8^. This suggests PSZ exhibit higher occupancy of D2Rs due to higher synaptic levels of DA, consistent with increased presynaptic dopamine release.

To investigate the molecular and circuit-level effects of dopamine dysregulation, a transgenic mouse model was developed with selective overexpression of D2Rs in the caudate-putamen^9^. These D2R-overexpressing (D2R-OE) mice display performance deficits on a working memory (WM) task that persist when D2R overexpression is halted in adulthood^9^, suggesting a neurodevelopmental cognitive deficit reminiscent of those seen in human PSZ^10, 11^. Notably, D2R-OE mice also display a neuroplastic alteration of projections in the basal ganglia-thalamo-cortical (BGTC) circuit^12^, which is broadly organized into two functionally opposed pathways (the “direct” and “indirect” pathways; see **Figure 1**) and plays a critical modulatory role in motor, limbic, and cognitive functioning^13, 14^. The BGTC circuit shows aberrant functional connectivity in PSZ at both the major input (striatal) and output (thalamic) nuclei of the circuit^15-25^, raising the possibility that alterations in this circuit observed in D2R-OE mice could have direct relevance for the neurobiology of schizophrenia.

**Figure 1.**
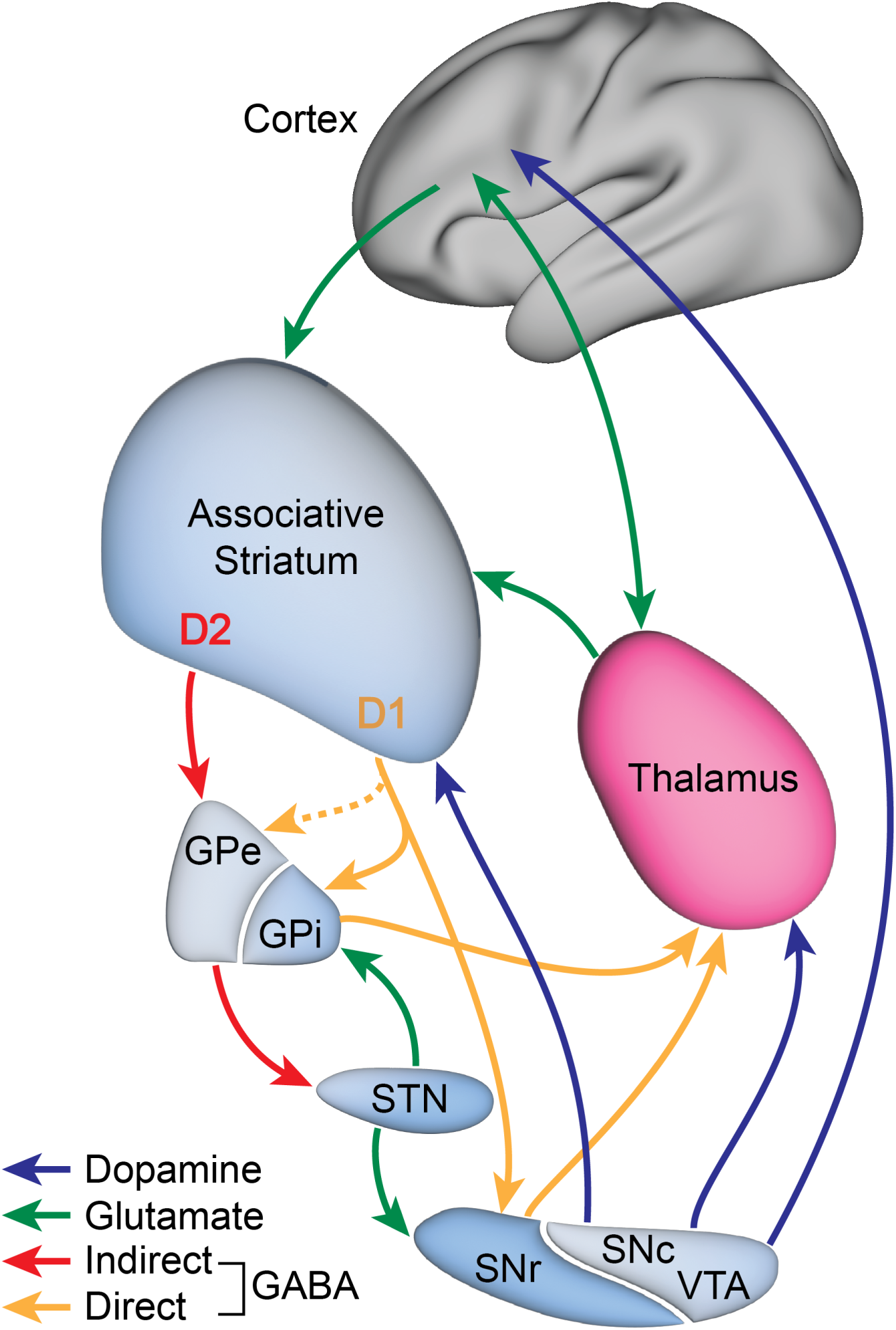
Schematic of basal ganglia-thalamo-cortical circuitry, highlighting glutamatergic (green arrows), GABAergic (red and orange arrows), and dopaminergic (blue arrows) connections between regions. Orange dashed line represents bridging collateral axonal projections from the associative striatum (primarily dorsal caudate) to the globus pallidus externus (GPe). GPi = globus pallidus internus; STN = subthalamic nucleus; SNr = substantia nigra pars reticulata; SNc = substantia nigra pars compacta; VTA = ventral tegmental area.

Specifically, D2R-OE mice exhibit increased density of “bridging collaterals” – axonal collaterals that branch from the globus pallidus internus (GPi)-projecting axons of striatal D1-containing MSNs and project to the globus pallidus externus (GPe)^12, 26-28^. Thus, these collaterals “bridge” the direct and indirect pathways. Critically, this increased density of bridging collaterals decreased when overexpression was halted in adult animals, but subsequently returned to elevated levels when D2R overexpression was restored. Haloperidol, a D2R antagonist, also reduces bridging collateral density in D2R-OE and wildtype animals, while D2R knockdown mice exhibit dose-dependent reductions in bridging collateral density^12^, demonstrating a causal relationship between striatal D2R stimulation and bridging collateral density. Notably, bridging collateral density in D2R-OE mice was greatest in associative striatum (medial caudate-putamen), which receives input from associative cortical regions such as the dorsolateral prefrontal cortex (DLPFC)^12^. Concordantly, PET imaging studies of PSZ have shown greater baseline occupancy of D2Rs in associative striatum, especially in the dorsal caudate (DCa)^7, 29^.

Here, we posit that if similar increased bridging collateral density occurs in human PSZ as a result of D2R overstimulation, it should be detectable as altered functional connectivity (FC) between the striatum (specifically DCa) and the GPe. However, evidence also suggests that: 1) any alteration in FC may only be observable in unmedicated PSZ, as D2R antagonists reduce bridging collateral density in rodents; 2) while altered anatomical connectivity should alter DCa-GPe FC, the expected direction of change is unclear—although striatal MSNs are GABA-ergic and thus inhibitory, net inhibition of a region is still metabolically demanding and can increase functional Magnetic Resonance Imaging (fMRI) Blood Oxygen Level-Dependent (BOLD) signal^30^; and 3) because striatal MSNs are hyperpolarized at rest, with low baseline firing rates and periodic bursting^31-34^, it is possible that altered FC may only be apparent under conditions in which the affected basal ganglia circuitry is actively processing information and MSNs are activated by cortical (and nigral) inputs.

Consequently, we conducted a resting-state and WM task-state FC MRI study (see, e.g., ^35, 36-45^) of unmedicated PSZ, using a WM task that robustly activates the DCa and other striatal regions^46^. We hypothesized that if bridging collateral density is increased in unmedicated PSZ, then DCa-GPe FC should be altered in PSZ, either during the performance of a WM task or both during WM task performance and at rest.

Given the interconnected nature of the BGTC circuit, we chose to implement a network modeling approach to calculating FC between regions (see, e.g., ^47^). This addresses the “third variable” problem, in which altered FC between regions could be driven by a third region that is not accounted for in the model (e.g., altered striato-pallidal connectivity could be driven by altered pallido-thalamic or cortico-striatal connectivity). Our approach calculates FC as partial correlations, controlling for timeseries in regions of interest (ROIs) outside each given ROI pair, a straightforward method that reliably distinguishes between direct and indirect network connections in simulated and *in vivo* data^47, 48^. Additionally, this approach partially accounts for partial volume effects arising in adjacent network structures (e.g., pallidal subdivisions) that could otherwise impact FC estimates.

Many of our participants were also enrolled in other neuroimaging studies, which included studies of dopamine function that could further inform whether altered FC might be driven by striatal dopamine dysfunction. Thus, in two subsets of our sample, we also report on the relationship between DCa-GPe FC and dopamine function measured via 1) neuromelanin-sensitive MRI (NM-MRI) of the SN (an indirect measure of long-term dopamine turnover that is correlated with psychotic symptoms; ^49, 50^) and; 2) Positron Emission Tomography (PET) with the D2/3 agonist radiotracer [^11^C]-(+)-PHNO^51, 52^ and an amphetamine challenge; however, the number of available PET participants was below generally accepted minimum sample sizes (n = 11 across both groups), and thus should be taken as supplemental and preliminary evidence that may support further investigation with appropriately powered samples. Finally, we conducted exploratory analyses examining the relationship between DCa-GPe connectivity and WM task performance and positive and negative symptoms in PSZ.

## 2. Materials and Methods

### 2.1. Overview

This is a case-control study of unmedicated PSZ and healthy controls (HC) aged 18-55 years old and demographically matched on age, parental socioeconomic status, race, and ethnicity. All procedures occurred at the New York State Psychiatric Institute (NYSPI) except PET scanning, which occurred at Yale University. PSZ were considered unmedicated if they were antipsychotic-naïve or antipsychotic-free for at least three weeks prior to recruitment. Additionally, antipsychotic-naïve participants had no prior use of long-acting injectable antipsychotics and less than 2 weeks of cumulative lifetime antipsychotic exposure. All participants were free of major neurological disorders, current substance use disorders, and psychiatric disorders other than schizophrenia, schizophreniform disorder, or schizoaffective disorder. PSZ were assessed for capacity to give informed consent using the MacArthur Competence Assessment Tool for Clinical Research^53^ prior to obtaining informed consent. Full inclusion and exclusion criteria are described in the **Supplementary Methods.** All procedures were approved by the NYSPI and Yale Institutional Review Boards. This study’s overall hypothesis, including the focus on the specific associative striatum/GPe subcircuit discussed here and use of task-state and resting-state FC and the network modeling analytic approach employed, were defined prior to enrollment and stated explicitly in the original NIH RePORTER abstract of the award that funded this work (1K01MH107763; see ^54^) alongside a separate, independent hypothesis reported elsewhere^15^, but was not formally pre-registered.

### 2.2. Participants

Thirty-seven PSZ and 30 HC completed RS fMRI, with a subset of 29 PSZ and 29 HC completing task-based fMRI. Additionally, 22 PSZ and 20 HC who performed task-based fMRI underwent NM-MRI. Finally, a smaller subset of task-based fMRI participants, 7 PSZ and 4 HC, underwent [^11^C]-(+)-PHNO PET with amphetamine challenge as part of a separate and unrelated study (currently unpublished). Participants completed several clinical and demographic assessments described in the **Supplementary Methods** and reported in **Table 1**. RS fMRI data and NM-MRI data from a subset of these participants has been reported elsewhere^15, 49^, while task-based fMRI data were acquired specifically for testing the *a priori* hypothesis presented in this manuscript, due to the concern that hyperpolarized striatal MSNs may not exhibit altered FC when not receiving substantial excitatory input from cortex.

**Table 1.**
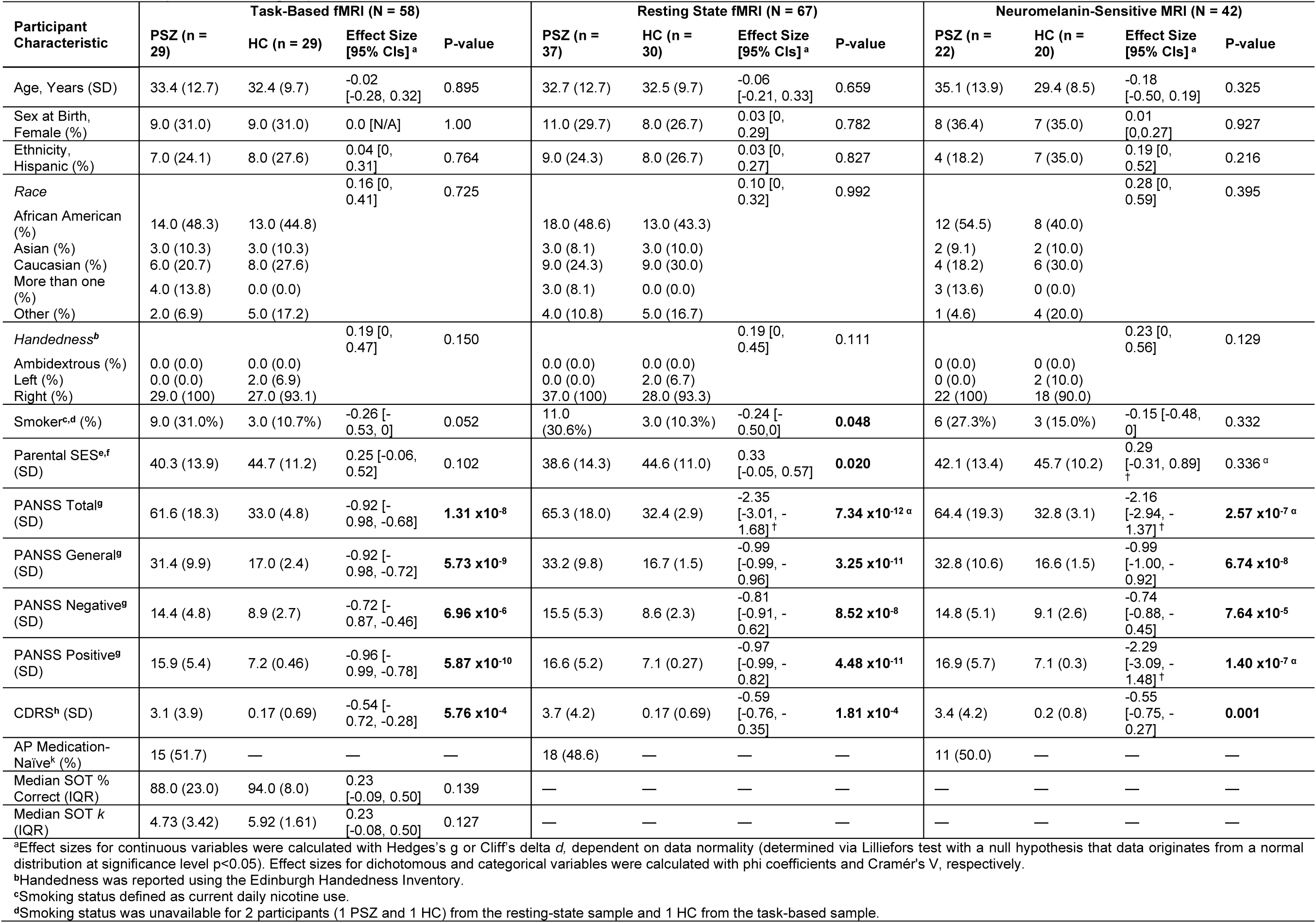
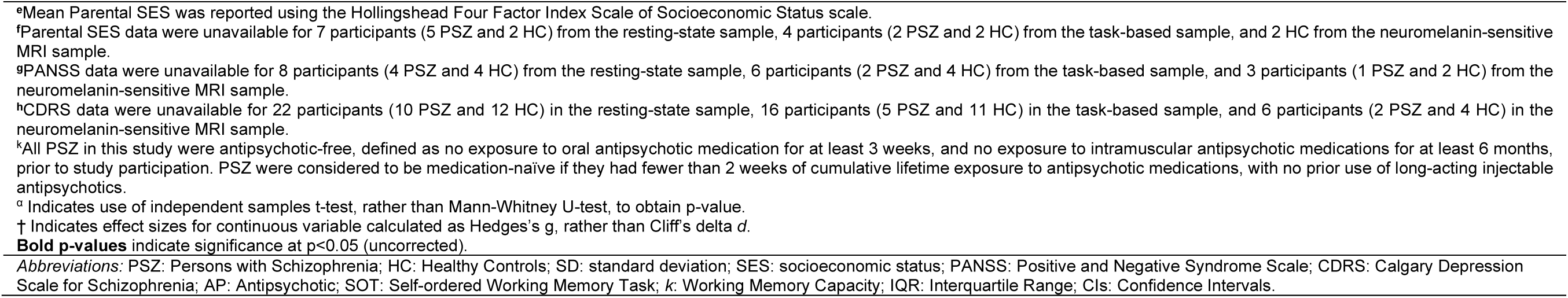
Participant Demographics and Clinical Measures.

### 2.3. Neuroimaging Procedures

MRI equipment, sequences, and processing are described in the **Supplementary Methods.** Briefly, in addition to standard structural sequences, we collected multiband fMRI scans with 2 mm isotropic voxel resolution for approximately 30 minutes each for RS and task-state FC (ts-FC). We employed the self-ordered WM task (SOT; see Figure S1 for task schematic; ^46, 55^); neural responses to task-events were modeled as per our prior work^46, 56^. WM task fMRI data were then residualized with respect to modeled task-evoked activation to distinguish ts-FC from task-related coactivation^57, 58^. Preprocessing was conducted with the Human Connectome Project Minimal Preprocessing Pipeline^59^, version 4.2.0, followed by established FC-specific processing methods, including data-driven motion censoring methods optimized for multiband fMRI in prior work^60^ (see **Supplementary Methods** for complete description). Finally, ts-FC used only “task-on” volumes, which were identified as volumes beginning two seconds after each task block onset and ending four seconds after the final trial of the block to account for the initial delay in hemodynamic response and gradual return to baseline^61^.

To use network modeling to estimate FC, we identified a set of regions in the BGTC circuit with established roles in cognitive and associative functions: the mediodorsal nucleus (MD), DCa, GPe, GPi, SN, and DLPFC (see **Supplementary Methods** for identification procedures, **Figures S2**, **S3** for ROI visualizations). We omitted the subthalamic nucleus from this model, despite its importance in the indirect pathway, due to its small size and the lack of a reliable localization technique. Note that subcortical ROI timeseries were extracted using participant-specific ROI definitions.

NM-MRI was acquired and processed as described elsewhere^49^ (see **Supplementary Methods**). Our NM-MRI outcome measure was the average contrast-to-noise ratio (CNR) extracted from *a priori* voxels within an overinclusive mask of the SN and ventral tegmental area (VTA), used in prior work^49, 50, 62^, whose CNR is correlated with both psychosis severity in PSZ (measured as PANSS Positive subscale score) and subclinical positive symptom severity in persons at clinical high risk for developing a psychotic disorder^49, 50, 63^.

PET data were acquired using procedures described elsewhere^64^; these procedures and analysis details are described in the **Supplementary Methods**.

### 2.4. Statistical Analysis

Effect sizes for group differences in continuous variables were calculated using Hedges’ *g* or Cliff’s delta *d* for normally and non-normally distributed samples (as determined via Lilliefors test), respectively. Effect sizes for categorical and dichotomous variables were calculated as Cramér’s *V* and phi coefficients, respectively, with p-values calculated using Chi-squared tests with significance level p<0.05. Group differences in continuous demographic variables and clinical assessments were assessed via independent samples t-tests or Mann-Whitney U-tests, depending on data normality. Group differences in SOT WM capacity *k*^55^ and proportion of correct responses, both between HC and all PSZ and between HC and medication-naïve and medication-free PSZ separately, were assessed using Mann-Whitney U-tests.

Group differences in ROI pair FC were assessed via independent samples t-tests or Mann-Whitney U-tests, depending on data normality. The primary hypothesis investigating group differences in DCa-GPe rs-FC and ts-FC was assessed for significance (two-tailed) at α=0.05 with Dunn–Šidák correction for the two comparisons testing our circuit-specific hypothesis arising from the D2R-OE mouse model. Post-hoc tests of group connectivity differences between all other ROI pairs were assessed separately at α=0.05 with false discovery rate (FDR) correction across all tests^65^, as we had no *a priori* hypotheses regarding connectivity amongst this set of regions. The differing choice of Dunn–Šidák and FDR for these two cases is due to the greater sensitivity of the former when only two tests are performed, while the latter has greater sensitivity for a larger number of tests. As a supplementary analysis, we assessed group differences in DCa-GPe ts-FC and rs-FC using a robust multivariable linear regression with a Huber weight function with age and sex at birth as covariates.

Associations between DCa-GPe ts-FC and SOT performance (measured as WM capacity k^55^), and average SN-VTA NM-MRI signal, were assessed via robust multivariable linear regression with a Huber weight function with age, sex at birth, and diagnosis as covariates; associations with PANSS Positive and Negative symptom scores were assessed in PSZ using the same covariates above except diagnosis. Supplementary analyses of the PET findings were conducted in identical models, using [11C]-(+)-PHNO baseline BPND and ΔBPND in place of NM-MRI signal. Associations with SN-VTA NM-MRI signal, baseline BP_ND_, and ΔBP_ND_ were FDR-corrected (α=0.05) as they constitute a single family of tests concerning dopamine tone and function. All betas are reported as standardized regression coefficients *β*\*.

Supplementary statistical analyses investigating effects of previous medication exposure in PSZ, smoking status as a covariate, diagnosis-by-connectivity interaction terms, and regression analyses performed in HC and PSZ separately, as well as repeated-measures ANCOVAs investigating modulation of between- and within-subject effects across both ts-FC and rs-FC, are described in the **Supplementary Methods**.

## 3. Results

### 3.1. Demographics

Thirty-seven PSZ and 30 HC completed at least two runs of resting-state fMRI (N=67), with zero participants excluded for excess motion. Additionally, 29 PSZ and 29 HC completed at least two runs of task-based fMRI (N=58), after excluding three PSZ and one HC due to excess motion. Summarized demographic information is shown in **Table 1** (see **Table S1** for demographics of the PET sample).

The median [IQR] SOT WM capacities (*k)* for HC and PSZ were 5.92 [1.61] and 4.73 [3.42], respectively (*P=*0.127; **Figure S4A**). The median [IQR] proportions of correct responses in HC and PSZ were 0.94 [0.08] and 0.88 [0.23], respectively (*P=*0.139; **Figure S4B**). In comparison to medication-free PSZ, medication-naïve PSZ displayed greater task performance (median [IQR] proportion correct of 0.83 [0.21] in medication-free and 0.97 [0.15] in medication-naïve, *P=*0.018) and SOT *k* (median [IQR] of 3.64 [4.60] in medication-free and 5.94 [2.90] in medication-naïve, *P=*0.031). SOT *k* and proportion of correct responses were greater in HC than medication-free PSZ (*P*=0.007 and *P=*0.003, respectively, **Figure S4C, D**), but not medication-naïve PSZ (*P*=0.902 and *P*=0.664, respectively).

### 3.2. Resting-state and Task-state Functional Connectivity

No significant group difference in DCa-GPe rs-FC was observed (median [IQR] of 0.05 [0.16] in HC and 0.08 [0.12] in PSZ, *P=*0.233; **Figure 2C**). In contrast, PSZ displayed significantly elevated DCa-GPe ts-FC relative to HC (mean [SD] of 0.05 [0.09] in HC and 0.11 [0.10] in PSZ, *P=*0.0252; **Figure 2D**). This pattern of significance between rs-FC and ts-FC remained when group differences were assessed via robust regression controlling for age and sex at birth (**Tables S2, 3**). The group difference in DCa-GPe ts-FC remained significant when additionally controlling for smoking status (**Table S4**), while no group difference in rs-FC was observed (**Table S5**).

**Figure 2.**
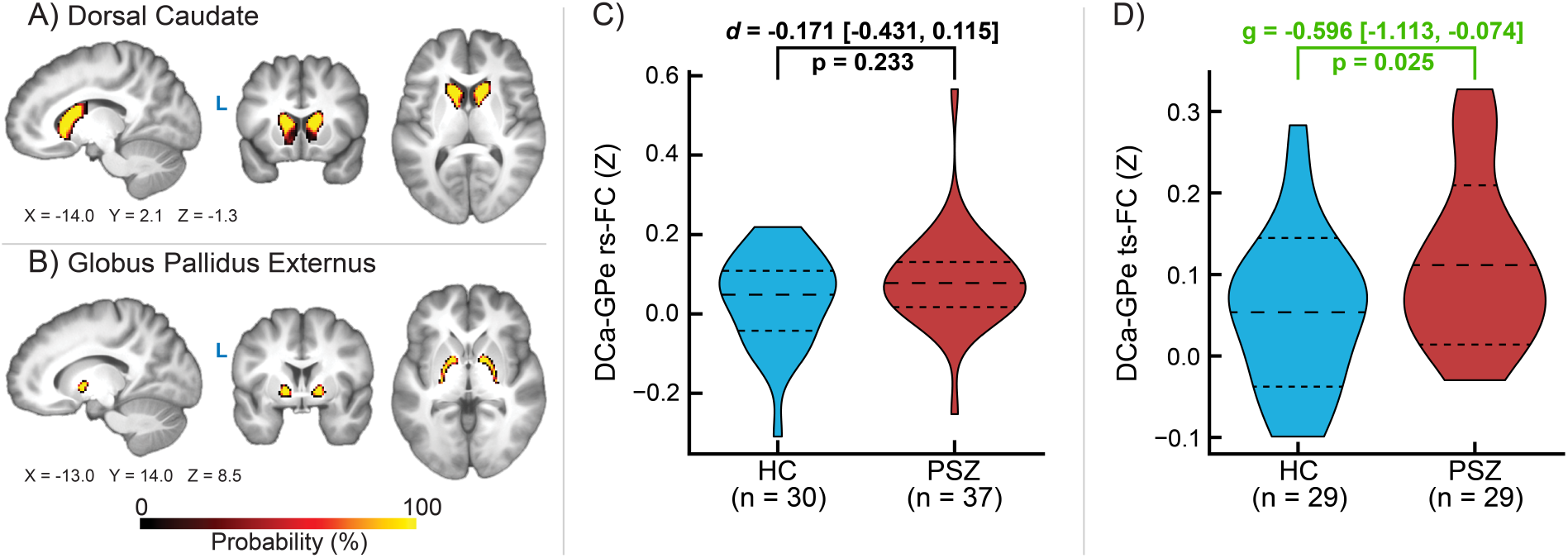
Probability density of **A)** dorsal caudate (DCa) and **B)** globus pallidus externus (GPe) regions of interest in the task-based fMRI sample (N = 58), as well as group differences between healthy controls (HC) and persons with schizophrenia (PSZ) in **C)** DCa-GPe resting-state functional connectivity (rs-FC) and **D)** DCa-GPe task-state functional connectivity (ts-FC). Long-dashed and short-dashed lines denote median and interquartile range in **Panel C** (due to non-normally distributed data) and means and standard deviations in **Panel D**. Both rs-FC and ts-FC were calculated while controlling for the average timeseries in dorsolateral prefrontal cortex, globus pallidus internus, substantia nigra, and mediodorsal nucleus. *g* = Hedges’ *g* [95% Confidence Intervals]; *d* = Cliff’s delta [95% Confidence Intervals].

No significant differences between medication-naïve and medication-free PSZ were observed in DCa-GPe ts-FC (mean [SD] of 0.12 [0.11] in medication-free and 0.10 [0.09] in medication-naïve, *P=*0.638) or rs-FC (median [IQR] of 0.07 [0.10] in medication-free and 0.08 [0.15] in medication-naïve, *P=*0.891). Additionally, no significant differences in DCa-GPe ts-FC or rs-FC were observed between HC and medication-naïve or medication-free PSZ when assessed separately (**Figure S5**), though both had comparable effect sizes to the primary group difference between HC and all PSZ, suggesting inadequate power for subgroup analyses.

Within other direct connections in the BGTC circuit, HC displayed greater MD-DCa rs-FC than PSZ (mean [SD] of 0.27 [0.12] in HC and 0.17 [0.15] in PSZ, *P=*0.005**; Figure S6H**). No significant differences in ts-FC were observed following FDR correction (**Figure S7**), although MD-DLPFC ts-FC was significantly elevated in PSZ before correction. Medication-naïve PSZ displayed lower DCa-SN rs-FC than HC (**Figure S8B**), and medication-free PSZ displayed lower MD-DCa rs-FC than HC (**Figure S8H**). No additional ts-FC differences were observed between HC and medication-naïve or medication-free PSZ separately after FDR correction (**Figure S9**).

### 3.3. Neuromelanin-Sensitive MRI

The average NM-MRI contrast-ratio map across participants is shown in **Figure 3A**. In *a priori* psychosis-associated voxels ^49, 50^, SN/VTA NM-MRI CNR was positively associated with DCa-GPe ts-FC (N=42; β* [SE]=0.40 [0.17], *P=*0.023; **Figure 3B**, **Table S6**) when controlling for age, sex at birth, and diagnosis, and survives FDR correction at α=0.05. This association remains significant when controlling for smoking status (**Table S7**). When investigating groups separately, the association remains significant in PSZ (N=22; β* [SE]=0.55 [0.22], *P=*0.019**; Table S8**) regardless of controlling for smoking status (**Table S9**). However, no significant interaction between diagnosis and ts-FC was observed (**Table S10**). Additionally, no significant effects of medication exposure or its interaction with DCa-GPe ts-FC were observed in PSZ (**Table S11**). Finally, a significant interaction between NM-MRI CNR and the connectivity type within-subject factor (ts-FC or rs-FC) in the repeated-measures ANCOVA was observed (**Table S12**); post hoc analysis revealed a stronger NM-MRI CNR relationship with DCa-GPe ts-FC than rs-FC.

**Figure 3.**
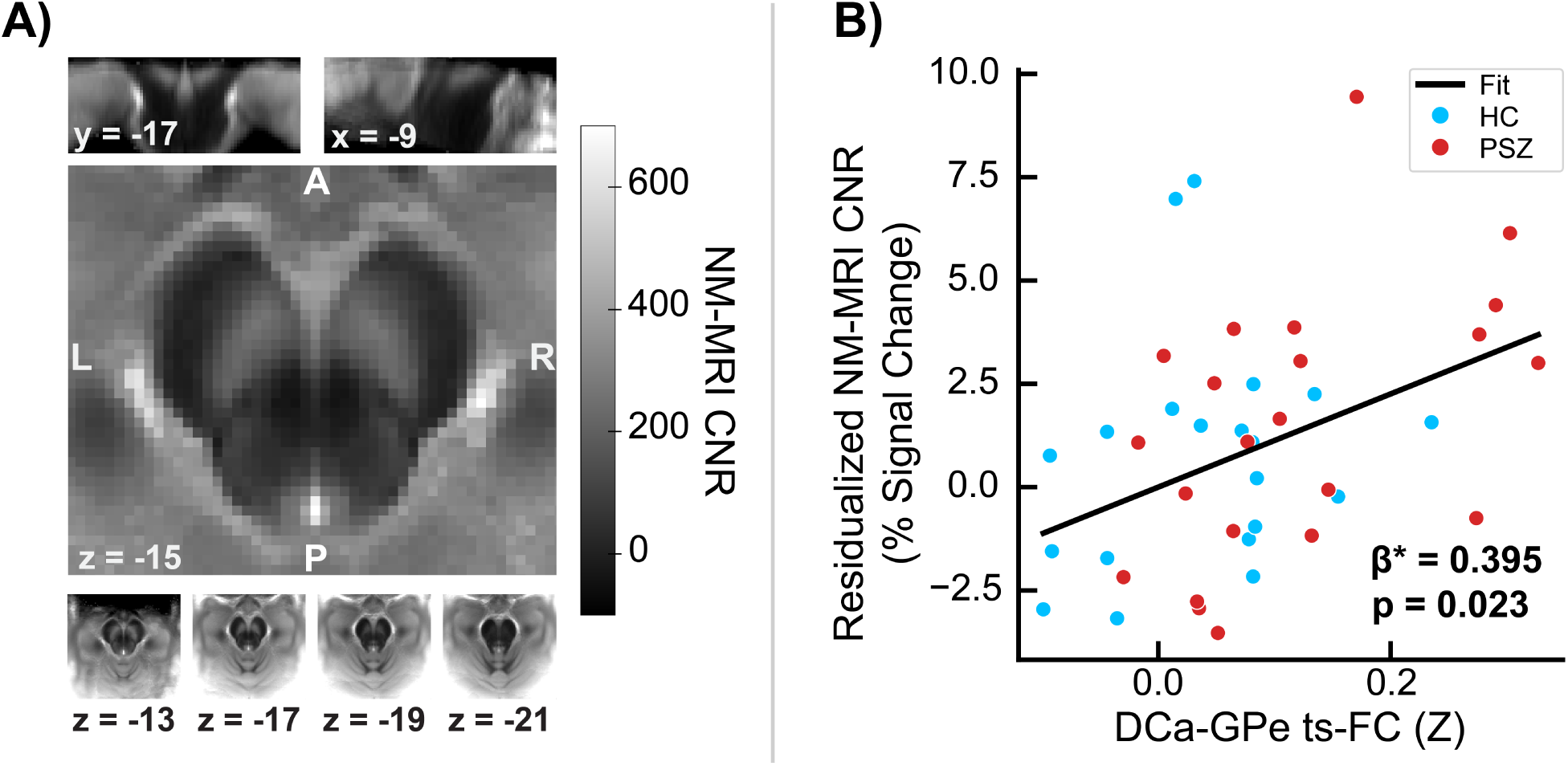
**A)** Average neuromelanin-sensitive MRI (NM-MRI) contrast-to-noise ratio (CNR) image across all participants (N = 42). **B)** Association between average substantia nigra – ventral tegmental area NM-MRI CNR and task-state functional connectivity (ts-FC) between dorsal caudate (DCa) and globus pallidus externus (GPe), controlling for age, sex at birth, and diagnosis (N = 42). Regression plot inlay shows standardized regression coefficient β*. Blue dots denote healthy controls; red dots denote persons with schizophrenia.

### 3.4. [^11^C]-(+)-PHNO Positron Emission Tomography

In the exploratory [^11^C]-(+)-PHNO PET sample, elevated DCa-GPe ts-FC was significantly negatively associated with DCa baseline D2R availability, measured as [^11^C]-(+)-PHNO BP_ND_ (N=11; β* [SE]=-0.45 [0.17], *P=*0.039; **Figure S10A, Table S13**). Additionally, greater DCa-GPe ts-FC was associated with more negative DCa ΔBP_ND_ (N=11; β* [SE]=-0.82 [0.27], *P=*0.021; **Figure S10B**, **Table S13**), where a more negative ΔBP_ND_ indicates greater fractional decrease in BP_ND_ following amphetamine administration. Both results survive FDR correction at α=0.05. After controlling for smoking status, there are no significant associations between DCa-GPe ts-FC and baseline BP_ND_ or ΔBP_ND_ (**Table S14**).

### 3.5. Associations between Task-state Functional Connectivity and Symptomatology

Elevated DCa-GPe ts-FC was associated with lower SOT *k* across all participants (N = 58; β* [SE]=-0.31 [0.13], *P=*0.020; **Figure 4A**, **Table S15**). This association remains significant when controlling for smoking status (**Table S16**) and within PSZ alone (N=29; β* [SE]=-0.36 [0.15], *P=*0.029; **Table S17**; see **Table S18** for inclusion of smoking status as a covariate), but not in HC. However, no significant diagnosis-by-connectivity interaction was observed (**Table S19**). Additionally, there was no significant interaction of medication exposure and DCa-GPe ts-FC in predicting SOT *k* (**Table S20**). Finally, a significant main effect of diagnosis and interaction between SOT *k* and the connectivity type within-subject factor (ts-FC or rs-FC) in the repeated-measures ANCOVA was observed (**Table S21**). Post hoc analysis indicated that PSZ possessed greater DCa-GPe connectivity across both connectivity conditions, and that the relationship between SOT *k* and connectivity was stronger for ts-FC than rs-FC.

**Figure 4.**
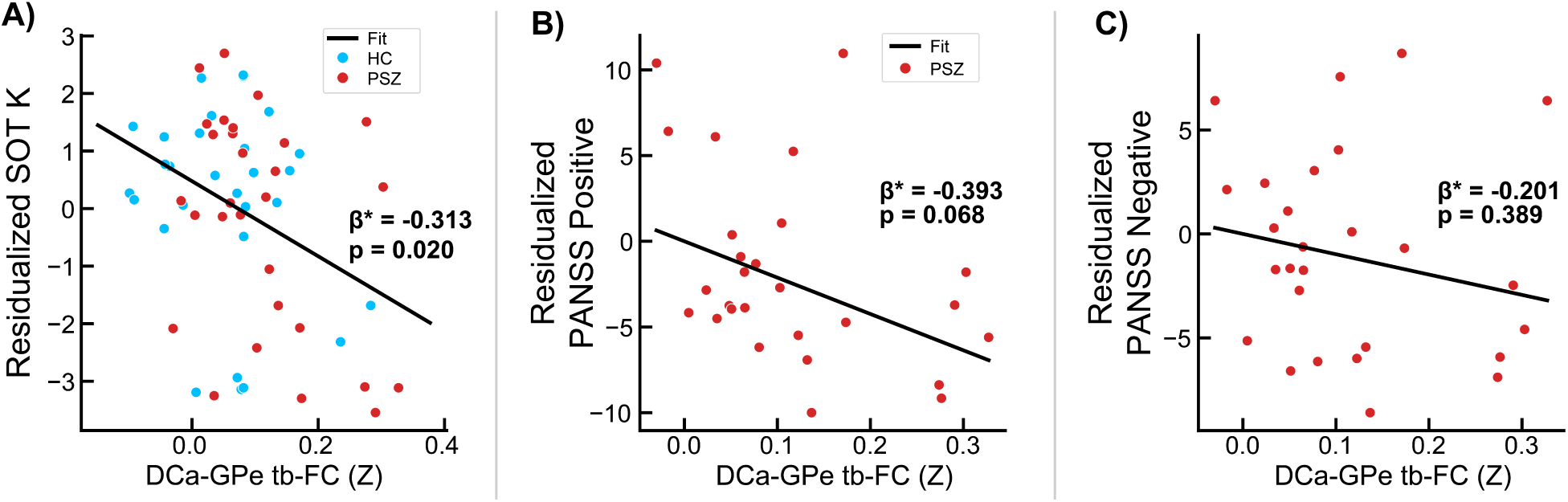
Associations between dorsal caudate-globus pallidus externus task-state functional connectivity (DCa-GPe ts-FC) and **A)** working memory capacity *k* calculated from self-ordered working memory task (SOT) performance, **B)** Positive and Negative Syndrome Scale (PANSS) Positive symptom score, and **C)** PANSS Negative symptom score. Associations between *k* and DCa-GPe ts-FC were calculated in all participants (N = 58), controlling for age, sex at birth, and diagnosis. PANSS symptom associations were calculated in persons with schizophrenia (PSZ; red) only (N = 27; PANSS data were unavailable for two PSZ), controlling for age and sex at birth. Plot inlays show standardized regression coefficient β*. Blue dots denote healthy controls (HC); red dots denote persons with schizophrenia (PSZ).

In PSZ, no significant associations were observed between DCa-GPe ts-FC and PANSS Positive symptom score (N=27; β* [SE]=-0.39 [0.21], *P=*0.068; **Figure 4B, Table S22**) or PANSS Negative symptom score (N=27; β* [SE]=-0.20 [0.23], *P=*0.389; **Figure 4C, Table S22**).

## 4. Discussion

The results presented here support our a priori hypothesis, defined prior to enrollment (see ^54^), that dorsal caudate-globus pallidus externus (DCa-GPe) connectivity would be specifically altered in unmedicated persons with schizophrenia, a prediction arising from reorganization of this circuitry observed in the D2R-OE mouse model^9, 12^ of striatal dopamine dysfunction in schizophrenia. Our findings also provide multiple lines of evidence that this alteration is associated with striatal dopamine dysfunction and WM deficits, consistent with dopaminergic and behavioral deficits observable in D2R-OE mice.

We demonstrated DCa-GPe hyperconnectivity in unmedicated PSZ that is observable during performance of a WM task, but not in the resting state. We anticipated that this may occur because MSNs are quiet at rest due to a low intrinsic resting membrane potential, which has been observed in *in vitro* human cells in addition to rodents and non-human primates^31-34^ (although the membrane potential and firing rate of MSNs has not been evaluated in PSZ with altered dopamine function), and may prevent detection of altered fMRI connectivity unless cortical and thalamic projections are actively stimulating striatal MSNs during cognitive processing. The results of secondary analyses are also consistent with our hypotheses, with significant correlations between DCa-GPe ts-FC and WM as well as all dopaminergic outcome measures collected in our sample (although, as noted, PET results should be considered preliminary given the small sample size), and these correlations with WM and NM-MRI were stronger for ts-FC than for rs-FC. Notably, DCa-GPe ts-FC was specifically elevated, rather than part of a broader alteration of ts-FC throughout the BGTC network (see **Figure S7**).

Consistent with evidence of dopaminergic control of bridging collateral density in D2R-OE mice, our findings suggest a potential role for subcortical dopamine dysfunction in altered striato-pallidal functional connectivity in PSZ. NM-MRI signal in the SN and VTA is a proxy measure of long-term dopamine turnover (unless neuronal cell death has occurred, e.g., in Parkinson’s disease^49, 50^) that was associated with psychotic symptom severity in prior work^66^. One potential source of elevated nigral NM signal is upregulated dopamine turnover in nigral cell bodies, which is suggested by PET studies of PSZ using [^18^F]DOPA^1, 2, 4^ and may result in elevated presynaptic availability and release in striatum^67^, a well-established finding in PSZ linked to psychosis severity^5, 7, 8, 29^. Although observed in a small exploratory sample of PSZ and HC, our finding that individuals who show stronger dopamine release following amphetamine challenge have more greatly elevated DCa-GPe connectivity provides limited preliminary support for this interpretation. Similarly, decreased baseline D2R availability was also associated with DCa-GPe hyperconnectivity in our PET sample, which could be due to increased baseline availability of dopamine in the DCa that competes with radiotracer binding, resulting in lower BP_ND_ in these participants. However, we cannot rule out the possibility that lower D2R density in DCa also contributes to lower BP_ND_.

Finally, we found that elevated DCa-GPe ts-FC is associated with poorer WM task performance; this association remains significant in PSZ alone and may mirror performance deficits in a T-maze WM task observed in D2R-OE mice^9^. Given that these deficits in D2R-OE mice remained when upregulation of DCa-GPe bridging collaterals was normalized via D2R antagonist antipsychotic administration, this finding raises the possibility that DCa-GPe hyperconnectivity could potentially be a state-dependent predictor of the neurodevelopmental impact of persistent dopamine dysfunction in PSZ prior to becoming medicated. That is, unmedicated PSZ with more severe subcortical dopamine dysfunction may develop greater bridging collateral density between DCa and GPe, driving further BGTC circuit reorganization that disrupts the maintenance of internal representations of task stimuli in prefrontal cortical regions, e.g., via effects on D1R function in DLPFC, which is thought to be disrupted in PSZ^68^, critical for WM, and altered in D2R-OE mice^9, 69^. Specifically, D2R-OE mice show persistent abnormalities in dopamine neurons, particularly decreased phasic activity and neuronal recruitment under WM task demands^70^. If these mechanisms are present in human PSZ, these abnormalities may not be normalized via antipsychotic medication administered after they develop, resulting in persistent WM deficits.

While these results provide support for our hypothesis of dopamine-driven alterations to DCa-GPe connectivity in PSZ, several caveats and limitations must be acknowledged. First, alternative interpretations of elevated DCa-GPe ts-FC in PSZ are plausible given that rs-fMRI and task-based fMRI cannot discern patterns of DCa-GPe structural connectivity, which should be further investigated in future work using diffusion weighted imaging. At the time the study was initiated, the resolution of diffusion imaging methods available to us was likely insufficient to resolve alterations in a short-distance tract between small basal ganglia structures, but FC changes in fMRI can reflect circuit-level consequences of structural changes that may not be resolvable with the older generation of diffusion imaging images.

Next, group differences in WM performance were only observed between HC and medication-free PSZ, not with medication-naïve PSZ. Indeed, medication-naïve PSZ performed slightly better on the SOT than HC, which suggests this sample represents a high-functioning, psychiatrically stable outpatient population. While this limits claims that can be made about the relevance of previous dopamine antagonist medication exposure to basal ganglia connectivity alterations explored in this study, we still observe comparable effect sizes in group differences in DCa-GPe ts-FC between both PSZ groups separately and HC (**Figure S5B**). Additionally, no significant medication exposure effects were observed in regressions predicting SOT *k* or SN-VTA NM-MRI CNR from DCa-GPe ts-FC, which remains a significant predictor of both outcomes when tested in PSZ alone. Nonetheless, effects of prior medication exposure cannot be completely ruled out in our sample.

In addition, limitations in sample size in both the RS fMRI and PET samples may limit the generalizability of the presented results. While larger than the task-based fMRI sample, the RS fMRI sample may have lacked statistical power to detect significant group differences in DCa-GPe connectivity at smaller effect sizes, although the observed difference trended in the expected direction. However, the RS fMRI sample did possess sufficient power to detect a larger effect in MD-DCa hypoconnectivity in PSZ (**Figure S6**), a finding consistent with previous work^71-73^. The PET sample should be considered extremely preliminary, as it was a very small sample of N = 11 with markedly younger PSZ than other samples in this study. Further, associations between PET metrics and DCa-GPe ts-FC did not survive when controlling for smoking status. Thus, associations between DCa baseline and ΔBP_ND_ and DCa-GPe ts-FC may not generalize to larger, more variable samples of PSZ. Finally, no significant associations were observed between DCa-GPe ts-FC and PANSS Positive score. While we expected DCa-GPe connectivity may be an overall marker of disease severity, the lack of significant association may arise from statistical power limitations. Alternatively, DCa-GPe connectivity may more closely index the severity of a phenotype that leads to WM deficits in PSZ, rather than psychotic symptoms. This proposition would ideally be studied in a larger, transdiagnostic sample with WM ts-FC.

In summary, this study provides *in vivo* translational evidence of a neural abnormality between the DCa and GPe in unmedicated PSZ, consistent with findings from the D2R-OE transgenic mouse model. This abnormality was observed as elevated FC during WM task engagement and was associated with poor WM task performance and altered subcortical dopaminergic function measured by two imaging modalities. Future work should aim to demonstrate altered DCa-GPe structural connectivity, as well as examine the role of D2R antagonist antipsychotic medication in the normalization of the phenotype. Nevertheless, these results build on a rich literature implicating subcortical dopaminergic dysfunction in the pathophysiology of schizophrenia to suggest a potential neurodevelopmental mechanism of WM deficits in PSZ, which could be a crucial step towards the development of novel therapeutic targets and treatments.

## 5. Data and Code Availability

All data and analysis code used in the preparation of this manuscript can be made available upon request to the corresponding author through a formal data sharing agreement.

## 6. Author Contributions

**Conceptualization:** J.X.V.S., A.A-D., C.K., and G.H.; **Data Curation:** P.N.T., J.C.W., Y.P., Z.J.Z., E.B.S-F., K.B., R.B.G., G.P., N.K.H., N.O., D.T.P., S.K.A., J.J.W., G.H., C.C., M.S., and J.X.V.S.; **Formal Analysis:** P.N.T., J.C.W., Y.P., D.T.P., Z.J.Z., M.S., C.C., and J.X.V.S.; **Funding Acquisition:** P.N.T., J.C.W., A.A-D., M.S., G.H., J.J.W., and J.X.V.S.; **Investigation:** P.N.T., J.C.W., Z.J.Z., R.B.G., E.B.S-F., K.B., G.P., D.T.P., N.O., N.K.H., S.K.A., J.J.W., M.S., A.A-D., G.H., C.C., and J.X.V.S.; **Methodology:** P.N.T., J.C.W., M.S., C.C., and J.X.V.S.; **Project Administration:** N.K.H., R.B.G., K.B., E.B.S-F., and J.X.V.S.; **Resources:** R.B.G., M.S., C.C., G.H., A.A-D., and J.X.V.S.; **Software:** P.N.T., J.C.W., S.K.A., Y.P., M.S., C.C., and J.X.V.S.; **Supervision:** R.B.G., N.K.H., G.P., J.J.W., M.S., G.H., C.C., A.A-D., and J.X.V.S.; **Validation:** P.N.T., J.C.W., Z.J.Z., Y.P., D.T.P., M.S., C.C., and J.X.V.S.; **Visualization:** P.N.T.; **Writing – Original Draft:** P.N.T., J.C.W., M.S., and J.X.V.S.; **Writing – Review & Editing:** P.N.T., J.C.W., G.P., G.H., A.A-D., C.K., M.S., C.C., and J.X.V.S.

## 8. Acknowledgements

The authors would like to thank Jaeyop Jeong, Sam R. Luceno, and Srineil Nizamabad for their contributions to fMRI preprocessing, Elizabeth Chan for assistance with data visualization, and Tram N.B. Nguyen and Dr. Benjamin A. Ely for helpful discussions. We would also like to acknowledge the substantial computing resources and technical assistance provided by Stony Brook Medicine Research Computing, with substantial support from Allen Zawada and James Xikis, as well as Stony Brook Research Computing and Cyberinfrastructure and the Institute for Advanced Computational Science at Stony Brook University for access to the high-performance SeaWulf computing system, with notable support from Fırat Coşkun, Daniel Wood, and David Carlson.

## Abbreviations

ANTs: Advanced Normalization Tools
BGTC: basal ganglia-thalamo-cortical
BOLD: blood-oxygen-level-dependent
BP_ND_: binding potential (non-displaceable)
CIFTI: connectivity informatics technology initiative
CNR: contrast-to-noise ratio
D1R: dopamine D1 receptor
D2R: dopamine D2 receptor
D2R-OE: dopamine D2 receptor overexpressing
DCa: dorsal caudate
DLPFC: dorsolateral prefrontal cortex
DOPA: dihydroxyphenylalanine
FC: functional connectivity
FDR: false discovery rate
fMRI: functional magnetic resonance imaging
GABA: gamma-aminobutyric acid
GEV-DV: generalized extreme value DVARS
GPe: globus pallidus externus
GPi: globus pallidus internus
HC: healthy control
HRF: hemodynamic response function
IBZM: iodobenzamide
IQR: interquartile range
MNI: Montreal Neurologic Institute
MP: motion parameter
MSN: Medium spiny neuron
NM-MRI: neuromelanin-sensitive magnetic resonance imaging
NYSPI: New York State Psychiatric Institute
PANSS: positive and negative syndrome scale
PET: positron emission tomography
PHNO: 4-propyl-9-hydroxynaphthoxazine
PSZ: persons with schizophrenia
ROI: region of interest
RS: resting-state
rs-FC: resting-state functional connectivity
SD: standard deviation
SE: standard error
SN: substantia nigra
SOT: self-ordered working memory task
SPM12: Statistical Parametric Mapping 12
STN: subthalamic nucleus
ts-FC: task-state functional connectivity
TE: echo time
TR: repetition time
VTA: ventral tegmental area
WM: working memory

