## Supplemental Information for "Translational Evidence for Dopaminergic Alteration of Basal Ganglia Functional Connectivity in Persons with Schizophrenia"

### Supplementary Methods

#### *Inclusion and Exclusion Criteria*

##### All Participants

Participants were recruited with advertisements, outpatient services at each institution, physician referrals, and via word-of-mouth. All individuals provided written consent prior to their participation and were free of psychoactive substances on the day of fMRI scanning as determined by a negative urine toxicology.

All participants were free of any major neurological disorders, current substance use disorders (≥3 months), and psychiatric disorders other than schizophrenia, schizophreniform disorder, or schizoaffective disorder. Psychiatric diagnoses were established according to the Diagnostic and Statistical Manual of Mental Disorders, Fourth Edition, Text Revision (DSM-IV-TR[1]), using the Structured Clinical Interview for the DSM-IV Axis I Disorders[2]. Clinical assessment scales were administered during initial screening by a PhD-level clinical psychologist or masters-level psychologist (GP or NO), and a detailed clinical history was obtained by a psychiatrist (RBG). Research procedures were initiated following a consensus diagnosis meeting.

All participants completed a magnetic resonance imaging (MRI) clearance form prior to any magnetic resonance (MR) scanning procedures to assess for the potential presence of metallic implants and past experiences with metal that could remain in the body. Participants were also administered a urine drug toxicology test before MR scanning procedures; in the event of a positive result for psychoactive drug use on the day of a scan, MR scanning was rescheduled at the discretion of participants and study personnel, provided the participant did not meet criteria for a substance use disorder. Participants assigned female at birth were administered a urine pregnancy screening test before MR scanning; in the event of a positive pregnancy test, the participant would be excluded from further participation in the study.

It is important to note that all participants were consented to complete both resting-state and task-based fMRI procedures. However, due to prioritization of the RS fMRI acquisition in the study protocol, as well as several participants terminating study participation prior to task-based fMRI acquisition, there are discrepancies in sample sizes in the resting-state and task-based samples utilized in this study (see **Table 1,** Methods *Section 2.2 Participants*).

**Healthy Control Participants (HC)**

Inclusion criteria for healthy control (HC) participants were: 1) ages 18-55, 2) absence of any current DSM-IV-TR (2) Axis-I psychiatric disorder, and 3) fluent in spoken English.

HC exclusion criteria were: 1) current substance use disorders; 2) current major affective episode; 3) history of neurological disorders, including head trauma with loss of consciousness, epilepsy, or other clinically significant disorders of the central nervous system, intellectual disability, hearing impairment, or unstable severe medical conditions; 4) claustrophobia; 5) metal implants or paramagnetic objects contained within the body; and 6) current pregnancy.

**Persons With Schizophrenia (PSZ)**

All persons with schizophrenia (PSZ) were either antipsychotic-free or antipsychotic-naïve for reasons unrelated to the study prior to their participation in the study. PSZ were considered to be antipsychotic medication-free if they had no exposure to oral antipsychotic medication for at least 3 weeks, and no exposure to intramuscular antipsychotic medications for at least 6 months, prior to their participation in this study. PSZ were additionally considered to be antipsychotic medication-naïve if they had fewer than 2 weeks of cumulative lifetime exposure to antipsychotic medications, with no prior use of long-acting injectable antipsychotics.

Inclusion criteria for PSZ were: 1) diagnosis of schizophrenia or schizoaffective disorder, per the DSM-IV-TR; 2) antipsychotic medication-free or antipsychotic-naïve (defined above); 3) age 18-55 years old, 4) fluent in spoken English; and 5) capacity to give informed consent, as determined using the MacArthur Competence Assessment Tool for Clinical Research.

Exclusion criteria for PSZ were: 1) current substance use disorders, 2) a current major affective episode, 3) history of neurological disorders, including head trauma with loss of consciousness, epilepsy, or other clinically significant disorders of the central nervous system, intellectual disability, hearing impairment, or unstable severe medical conditions, 4) claustrophobia; 5) metal implants or paramagnetic objects contained within the body; and 6) current pregnancy.

***Clinical Assessments***

Symptom severity was assessed for PSZ and HC using Positive and Negative Syndrome Scale (PANSS)[3], and for a subset of participants using the Psychotic Symptom Rating Scales (PSYRATS)[4], Scales for the Assessment of Negative Symptoms (SANS)[5, 6], and Calgary Depression Scale for Schizophrenia (CDRS)[7, 8]. Note that only PANSS symptom sub-scores were used in subsequent analyses. Handedness was assessed using the Edinburgh Handedness Inventory (EHI)[9]. Socioeconomic status (SES) was determined using the Hollingshead Four Factor Index of Socioeconomic Status[10]. Family psychiatric history was assessed using the Family History Screen (FHS)[11]. Given incomplete data collection in many assessments (primarily due to time constraints in participant study visits; see footnote of **Table 1**), symptomatology analyses were limited to the use of positive and negative symptom subscales from the (PANSS).

***Magnetic Resonance Imaging Acquisition and Processing***

Magnetic resonance imaging (MRI) was completed using a 3T General Electric (Boston, MA) MR750 with a NOVA 32-channel head coil (Nova Medical, Inc., Wilmington, MA). fMRI data were acquired with TR=850 ms, TE=25 ms, 192x192 mm field of view, and multiband factor of 6. Each resting-state (RS) fMRI run comprised 525 volumes (7.5 min) and each task-based run comprised 536 volumes (7 min 35 s per run). Runs were acquired using alternating anterior-posterior and posterior-anterior phase-encode directions. In addition to RS and task-based fMRI, the following were also acquired in each session: 1) T1-weighted and T2-weighted structural scans with 0.80 mm isotropic voxels, 2) spin-echo field maps in anterior-posterior and posterior-anterior phase-encode directions, and 3) a B0 field map.

RS and task-based fMRI were processed using the Human Connectome Project Minimal Preprocessing Pipelines, version 4.2.0[12]. Briefly, data underwent processing through the *PreFreeSurfer, FreeSurfer, PostFreeSurfer, fMRIVolume,* and *fMRISurface* pipelines. Volumetric images were normalized to Montreal Neurological Institute (MNI) 152 non-linear 6th-generation space [13], and surface images were obtained in CIFTI (Connectivity Informatics Technology Initiative) format. All data post-processing and statistical analysis following preprocessing was performed using custom analysis scripts in MATLAB (Natick, MA) R2023b.

***Self-Ordered Working Memory Task Procedures***

The Self-ordered Working Memory Task (SOT) was described in previously published work[14-16], and consisted of 20 trials, with eight steps of gradually increasing working memory (WM) load in each trial. At the start of a trial, a three-by-three grid of eight simple line drawings of three-dimensional objects was presented, with the center position in the grid left blank. Unique stimuli were used on each of the first 10 trials (80 total stimuli) and were each repeated once in the following 10 trials. Participants were given seven seconds in which to respond on each step. Responses consisted of using an fMRI compatible trackball (Current Designs, Inc., Philadelphia, PA) to position a cursor over one of the objects and select it with a button press.

Participants were instructed to select any object on each step that they had not already selected on a previous step (thus, on the first step all possible responses are correct). Once participants made a selection, a white square was displayed around the selected object until a total of nine seconds had elapsed since the beginning of the step, thus ensuring that each step remained the same length regardless of participants' reaction times. At the start of each subsequent step after the first, the objects were pseudo-randomly rearranged in the grid, but with the blank space placed at the location of the previously selected item (thus preventing participants from simply selecting the same location on each step). If participants failed to make a response within seven seconds from the beginning of a step, a white square was displayed around a randomly selected object that would have been a correct response. Participants were instructed to remember this object as if they had selected it themselves, and to continue the trial. If an incorrect selection was made, a red square was displayed over top of the selected object in order to indicate that an error had been made, and the same procedure as in the case of no response was followed. At the conclusion of each trial, an eight second pause occurred prior to the beginning of the next trial. Participants also carried out two trials of a control task occurring in the second and third task runs, in which one of the objects on each step was marked with an asterisk and participants were instructed to simply select the marked object. In all other respects the display and randomization of stimuli for the control task was identical to the SOT. Unique stimuli were used for each of the control trials, and each trial of either type was preceded by textual instructions indicating whether the upcoming trial was a task trial or a control trial. Task performance was measured as each participant's WM capacity *k*, with its calculation detailed elsewhere[14]. **Figure S1** shows a schematic of the first three steps of one trial.

***Self-Ordered Working Memory Task Modeling***

Within-participant task modeling was performed using SPM12[17, 18]. The SOT was modeled using an event-related design, with separate regressors of interest for correct responses on the control condition and each of the eight steps. Additional regressors were included for incorrect responses to control trials, incorrect responses to any step trial, instruction text, and button presses (to control for the influence of motor responses on estimates of task-evoked activation). All regressors described above were convolved with a three-parameter hemodynamic response function (HRF), with parameters for the canonical HRF plus the temporal and dispersion derivatives[15]. Nuisance regressors (which are not convolved with the HRF) include six MPs, their squares, derivatives, squared derivatives, and spike regressors reflecting volumes of high motion, as identified using run-adaptive, generalized extreme value, low-pass filtered DVARS (GEV-DV) thresholds[19].

***Task-state and Resting-state Functional Connectivity Processing***

Following WM task linear modeling, estimates of task activation for each run were obtained via robust linear regression with Huber weighting, and all task-based data was residualized with respect to only the regressors convolved with a three-parameter hemodynamic response function in the task design matrix (i.e., motion effects and spike regressors were modeled but not removed) to distinguish task-state FC (ts-FC) from task-related coactivation[20, 21]. Each run was then mode-1000 normalized[22, 23], linearly detrended, and mean-centered. Volumes acquired during periods of excess participant motion were removed using volume censoring with study-wide motion thresholds, measured by low-pass-filtered framewise displacement (LPF-FD), and run-wise thresholds for whole-brain signal fluctuation, using GEV-DV thresholding (see below, *Volume Censoring*)[19]. Censoring thresholds were calculated using only “task-on” volumes, which were identified as all volumes beginning two seconds after each task block onset and ending four seconds after the final trial of the block; this accounts for the initial delay in hemodynamic response and gradual return to baseline[24]. Runs were then filtered using a 0.009-0.08 Hz band-pass second-order zero-phase Butterworth filter, with censored time points replaced by linear interpolation prior to band-pass filtering before being discarded from analysis.

The first and last 22 seconds of each timeseries were discarded to remove discontinuity artifacts present after filtering.

RS runs were processed identically, with the exception of 1) no regression of task activation, and 2) volume censoring thresholds were estimated from all volumes, rather than just “task-on” volumes.

***Volume Censoring***

Volumes acquired during periods of excess participant motion were removed using volume censoring with study-wide motion thresholds, measured by low-pass-filtered framewise displacement (LPF-FD), and run-wise thresholds for whole-brain signal fluctuation, using GEV-DV thresholding[19]. Thresholds were determined in a data-driven manner for task-based and resting-state data separately using the Multiband Censoring Optimization Tool (MCOT)[19]. In resting-state runs, only, volumes acquired during eye closures lasting longer than 3 seconds were censored; volumes acquired between censored eye closures were additionally themselves censored if the time separating censored eye closure periods was less than 30 seconds. In all runs, contiguous clusters of data shorter than 8 s in duration after censoring were removed; any runs with less than 1.5 minutes of remaining data were subsequently discarded. After censoring in either task, participants with fewer than 2 runs of data, or fewer than 5 minutes of data total, were excluded from further analyses for that task. For resting-state data, the thresholds identified for LPF-FD and GEV-DV were 98.15 mm and 8.9 (arbitrary units), respectively. For task-based data, thresholds for LPF-FD and GEV-DV were 62.86 mm and 5.7 (arbitrary units), respectively.

***fMRI Regions of Interest***

Mediodorsal nucleus (MD) was identified using an automatic segmentation based on a probabilistic atlas from 12 postmortem thalami developed by Iglesias and colleagues[25]. The union of parvocellular and magnocellular MD subdivisions identified by the segmentation algorithm was used in order to ensure that the region of interest (ROI) signal was estimated from a sufficient number of voxels in each participant. The DCa ROI was taken as the precommissural caudate ROI generated via a deep-learning based segmentation algorithm trained on 68 hand-drawn striatal segmentations[26], which were drawn based on operational criteria established in our prior work[27, 28]. The GPe, GPi, and SN ROIs were extracted using DBSegment[29], a segmentation algorithm based on a nnU-Net[30] convolutional neural network trained on three prior atlases[31-33]. The SN ROI was taken as the union of the SN pars reticulata and pars compacta ROIs from DBSegment, again to ensure that signal was estimated from a sufficient number of voxels for each participant. It is important to note that all volumetric ROIs were generated in each participant's native T1 space, and subsequently warped to MNI space and resampled to 2mm resolution via enclosing voxel interpolation in Connectome Workbench v1.5[34] prior to timeseries extraction. Bilateral DLPFC ROIs were derived from a group-level contrast reflecting increased task-evoked activation relative to the control condition in healthy participants performing the SOT in prior work[16]; each ROI was taken as the largest contiguous DLPFC cluster after dilating by 4mm. DLPFC was chosen as a representative cortical region for the circuit under investigation due to its robust activation to SOT task demands[15, 16]. All ROI timeseries were extracted from spatially unsmoothed volumetric data with the exception of DLPFC, which was extracted from surface-based data.

***Functional Connectivity Analysis***

ROI pair rs-FC and ts-FC were calculated as the partial Pearson correlation between ROI timeseries, controlling for the following nuisance parameters: band-pass filtered motion parameters (MPs; using the same 0.009-0.08 Hz band-pass filter that was applied to each timeseries), the squares, derivatives, and squared derivatives of the band-pass filtered MPs, the white matter signal and its derivative, cerebrospinal fluid signal and its derivative, and the global signal and its derivative. Ts-FC was calculated using only "task-on" volumes identified previously (see above, *Task-state and Resting-state Functional Connectivity Processing*). Additionally, FC was calculated while controlling for all other ipsilateral ROI timeseries outside of those in the ROI pair as nuisance parameters. Final measures of ROI pair FC were calculated as the average of both ipsilateral ROI pairs, calculated separately. All ts-FC and rs-FC values were Fisher's r-to-Z transformed prior to averaging.

***Neuromelanin-Sensitive MRI Acquisition and Processing***

For a subset of participants, a single volume of NM-MRI data was acquired using a two-dimensional gradient recalled echo sequence with magnetization transfer pulse [35], providing partial brain coverage around the midbrain and pons, TR=260 ms, TE=2.68 ms, flip angle=40°, magnetization transfer frequency offset = 1,200 Hz, 0.43 mm × 0.43 mm in-plane resolution, 3 mm slice thickness, 10 slices, and five total excitations (averaged online to produce a single volume).

NM-MRI data was processed according to procedures introduced elsewhere[36]. First, each participant's NM-MRI scan was normalized to MNI space using Advanced Normalization Tools (ANTS) Version 2.5.3[37-40]. Contrast-to-noise ratio (CNR) for each voxel, *v*, was then calculated as the relative difference in NM-MRI signal intensity, *I*, from a reference region, *RR*, such that $CNR_{\mathrm{v}} = (I_v - mode(I_{RR}))/mode(I_{RR})$. The reference region used was a crus cerebri mask, a white matter tract with negligible neuromelanin content, obtained from prior work[36]. Images were then smoothed with a 1mm full-width-at-half-maximum gaussian kernel using SPM12[17, 18].

***[$^{11}$C]-(+)-PHNO Positron Emission Tomography Acquisition and Processing***

Following acquisition, PET data were reconstructed by the MOLAR (Motion-compensation OSEM List-mode Algorithm for Resolution-recovery reconstruction) algorithm[41]. Participants were scanned with the dopamine D3-preferring D2/D3 agonist radiotracer [$^{11}$C]-(+)-PHNO. Emission data were acquired for 120 min following a slow bolus (5 min) of radiotracer (307 ± 84 MBq, 1.51 ± 0.53 µg cold mass). A baseline scan was performed followed immediately by administration of oral d-amphetamine (0.5 mg/kg). A second scan was performed 3 hours after amphetamine administration. Participants' T1-weighted MRI scans were reoriented coronally in alignment with the anterior commissure-posterior commissure plain. PET data were coregistered to the MRI and ROIs were drawn manually on each participant's MRI and applied to the dynamic PET data to generate time activity curves. For this study, ROIs included DCa and cerebellum as a reference tissue. Kinetic analysis was performed using a basis function version of the simplified reference tissue model[42, 43] to generate the primary outcome measure binding potential $BP_{ND}$[44]. Capacity for dopamine release was inferred from the relative change in $BP_{ND}$ between conditions, $\Delta BP_{ND} = \frac{BP_{ND}(post-amph)}{BP_{ND}(baselline)} - 1$. In this definition, a more negative $\Delta BP_{ND}$ denotes greater post-amphetamine dopamine release.

Associations between DCa-GPe ts-FC and [$^{11}$C]-(+)-PHNO baseline $BP_{ND}$ and $\Delta BP_{ND}$ calculated within a hand-drawn DCa ROI were assessed via robust multivariable linear regression with a Huber weight function with age, sex at birth, and diagnosis as covariates.

***Supplementary Statistical Analyses***

In addition to group differences in ts-FC and rs-FC between HC and all PSZ, we additionally investigated differences in FC between HC and medication-free and medication-naïve PSZ separately. False discovery rate (FDR) correction was applied within the ts-FC and rs-FC families of comparisons. Next, regressions predicting SOT performance and SN-VTA NM-MRI were repeated in HC and PSZ, separately; within PSZ, analyses were repeated again with medication exposure and the interaction between DCa-GPe ts-FC and medication exposure as additional predictors. In addition, differences in slope between groups in the above regressions were assessed via diagnosis-by-connectivity or diagnosis-by-NM CNR interaction terms, respectively.

Further, group differences in DCa-GPe ts-FC and rs-FC, as well as associations between FC and NM-MRI signal, [$^{11}$C]-(+)-PHNO PET metrics, and SOT performance were repeated with smoking status included as a covariate; nicotine is a known modulator of midbrain dopamine function[45].

Finally, we performed repeated-measures ANCOVAs assessing whether the relationship between connectivity and either SOT *k* or NM CNR is dependent on the nature of the connectivity (DCa-GPe ts-FC or rs-FC). In each ANCOVA, between-subject factors comprised diagnosis and either SOT *k* or NM CNR, respectively, while the within-subject factor was connectivity type; age and sex at birth were included as covariates. Models included a three-way interaction term between diagnosis, connectivity type, and either SOT *k* or NM CNR. Each effect was assessed via F statistic, and partial eta-squared was calculated as a measure of effect size.

### Supplementary Figures and Tables

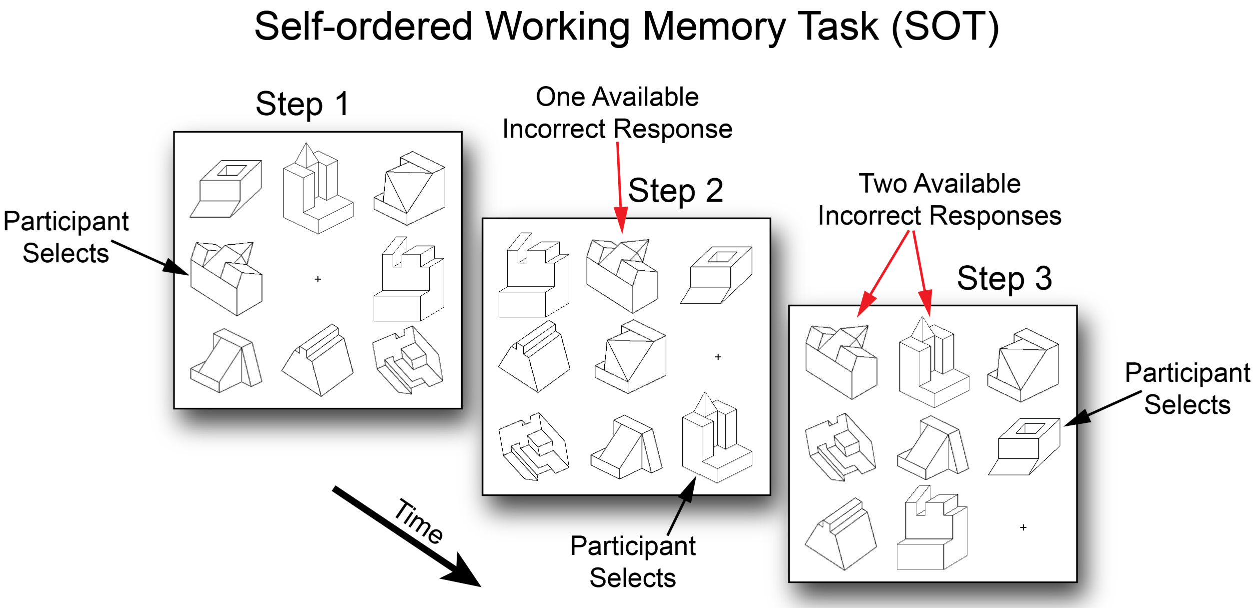


**Figure S1.** Schematic of the first three of eight steps in an 8-item self-ordered working memory task (SOT). At Step 1, participants may select any object. At Step 2, participants may select any object other than the that was selected on the prior step. At Step 3, any object other than the two previously selected objects may be selected. This proceeds until the participant has made 8 selections.

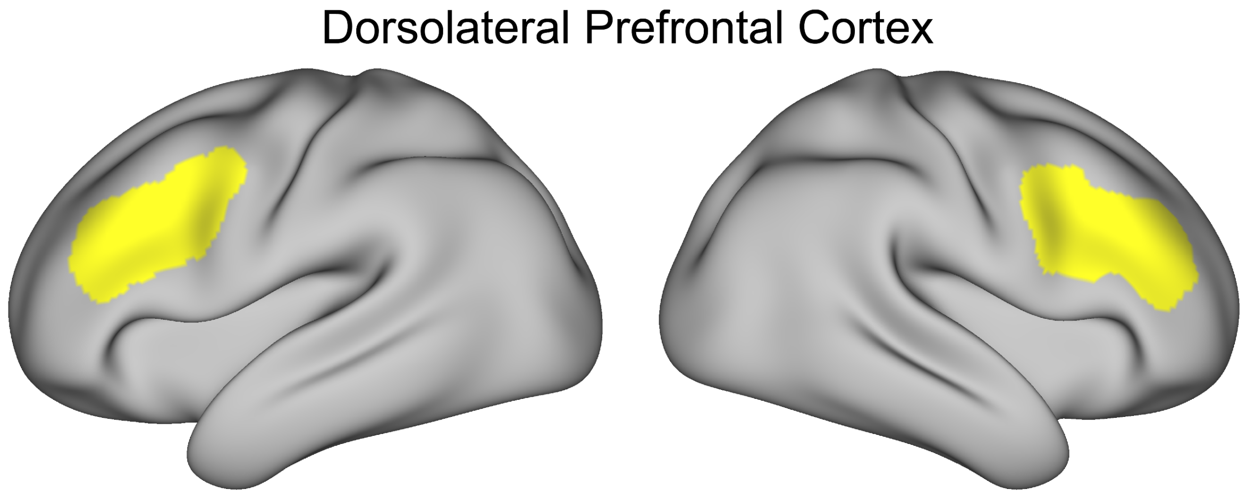


**Figure S2.** Dorsolateral prefrontal cortex region of interest used in this study, overlaid on the Human Connectome Project 1200 group average inflated surface.

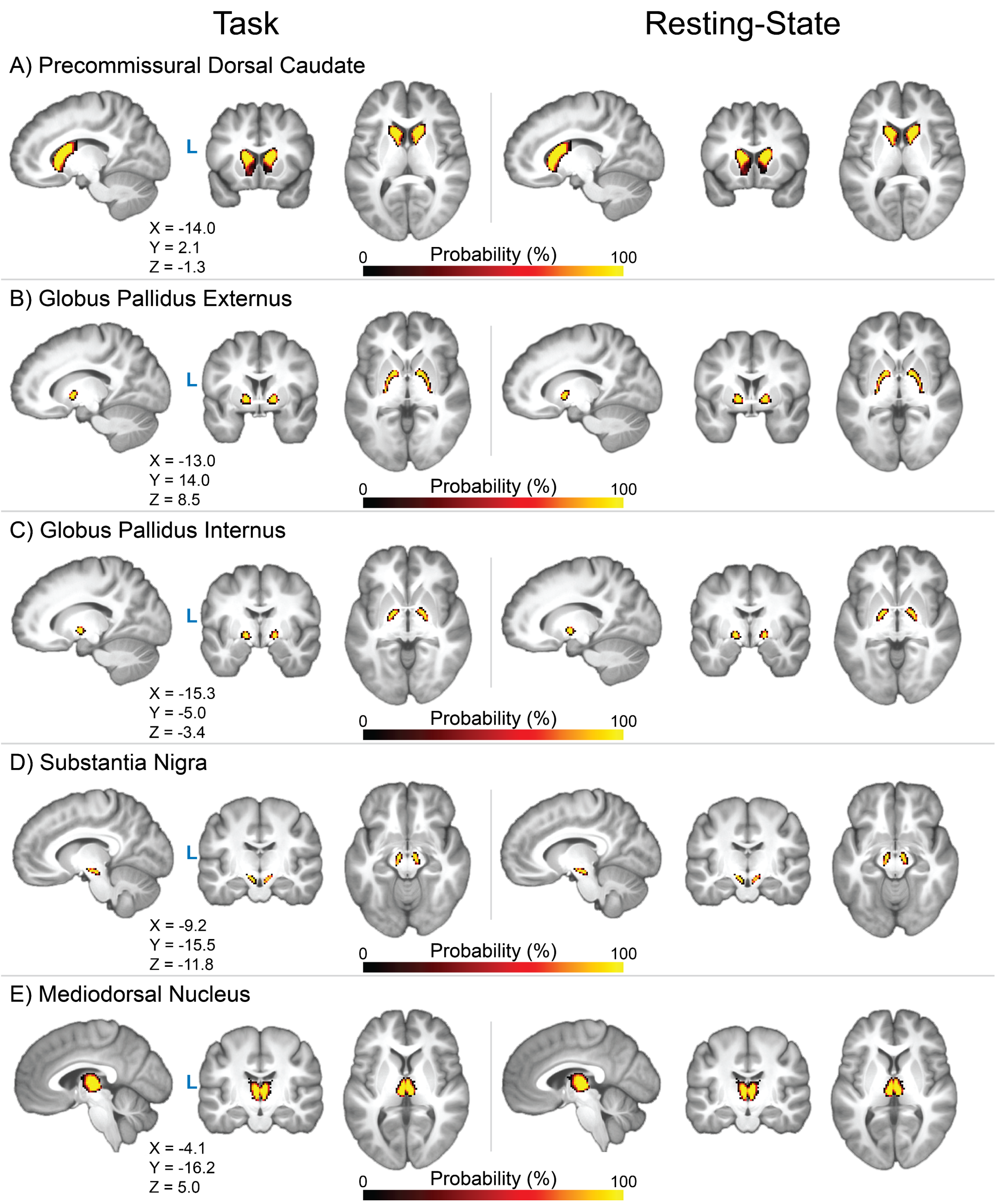


**Figure S3.** Probability density maps of volumetric regions of interest (ROIs) defined in this study for the task-based (left column; N = 58) and resting-state (right column, N = 67) fMRI samples. X, Y, and Z MNI152NLin6 coordinates are shown for each view. Regions of interest are overlaid on group average T1-weighted images for each sample. Note that ROI timeseries were extracted using participant-specific ROI definitions, not entire regions of probability density shown here across participants.

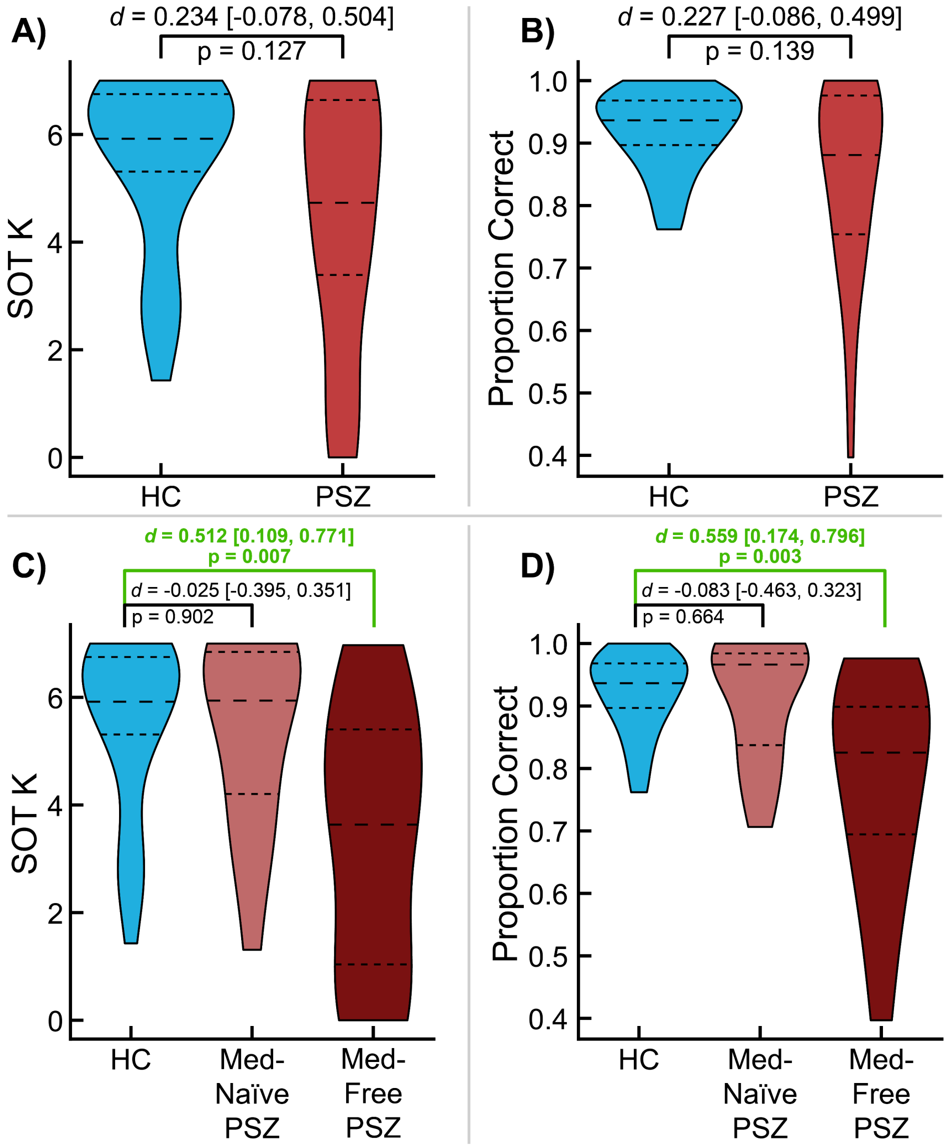


**Figure S4.** Group differences in working memory capacity *k* (Panel A), and proportion of correct responses (Panel B) in the self-ordered working memory task (SOT) between healthy controls (HC; blue) and persons with schizophrenia (PSZ; red). Differences in *k* (Panel C) and proportion of correct responses (Panel D) were additionally assessed between HC and medication-naïve and medication-free PSZ, respectively. Green text denotes statistical comparisons that survive false discovery rate correction across all comparisons presented in Panels C and D. Long-dashed and short-dashed lines denote medians and quartiles, respectively. *d* = Cliff's delta [95% Confidence Intervals].

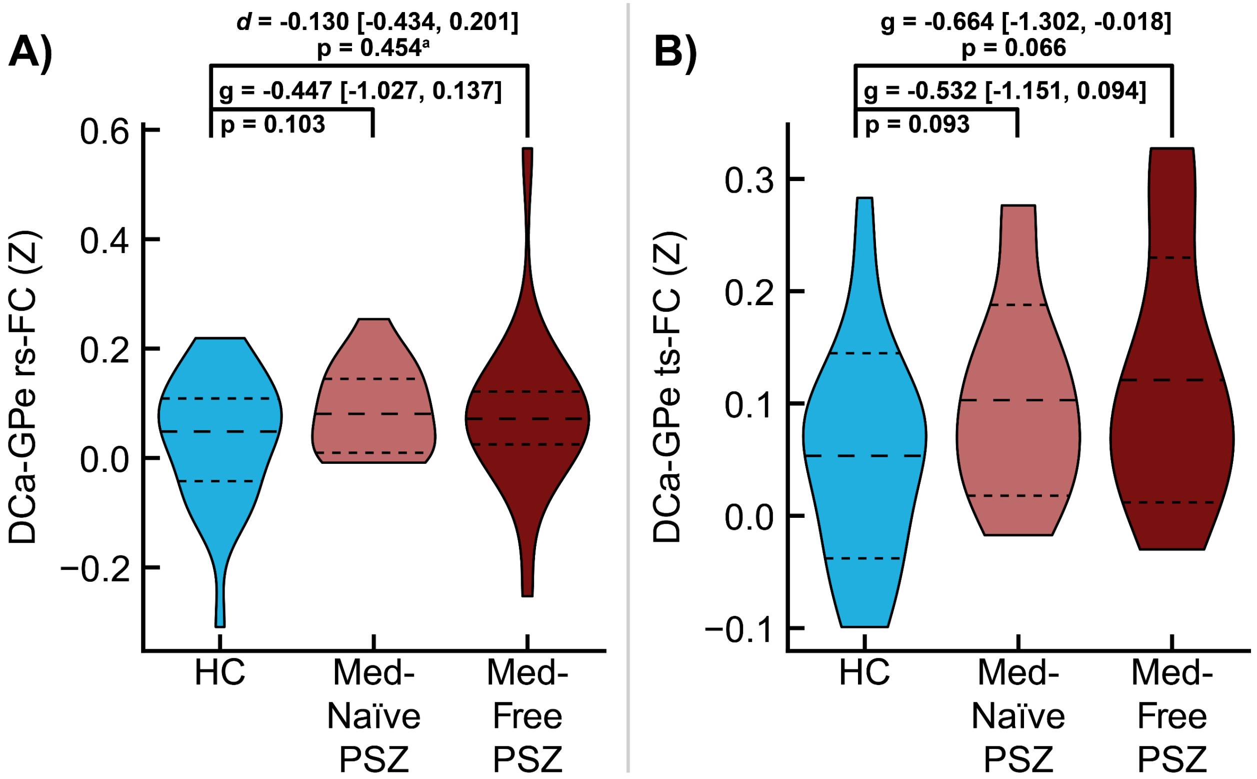


**Figure S5.** Group differences between healthy controls (HC) and medication-naïve and medication-free persons with schizophrenia (PSZ), respectively, in **C)** DCa-GPe resting-state functional connectivity (rs-FC) and **D)** DCa-GPe task-state functional connectivity (ts-FC). Long-dashed and short-dashed lines denote median and interquartile range in **Panel C** (due to non-normally distributed data) and means and standard deviations in **Panel D**. Larger outer brackets denote comparisons between HC and medication-free PSZ, while smaller inner brackets denote comparisons between HC and medication-naïve PSZ. Both rs-FC and ts-FC were calculated while controlling for the average timeseries in dorsolateral prefrontal cortex, globus pallidus internus, substantia nigra, and mediodorsal nucleus. *g* = Hedges' *g* [95% Confidence Intervals]; d = Cliff's delta [95% Confidence Intervals].

[a]Statistical testing was performed using Mann-Whitney U-test, rather than independent samples T-test.

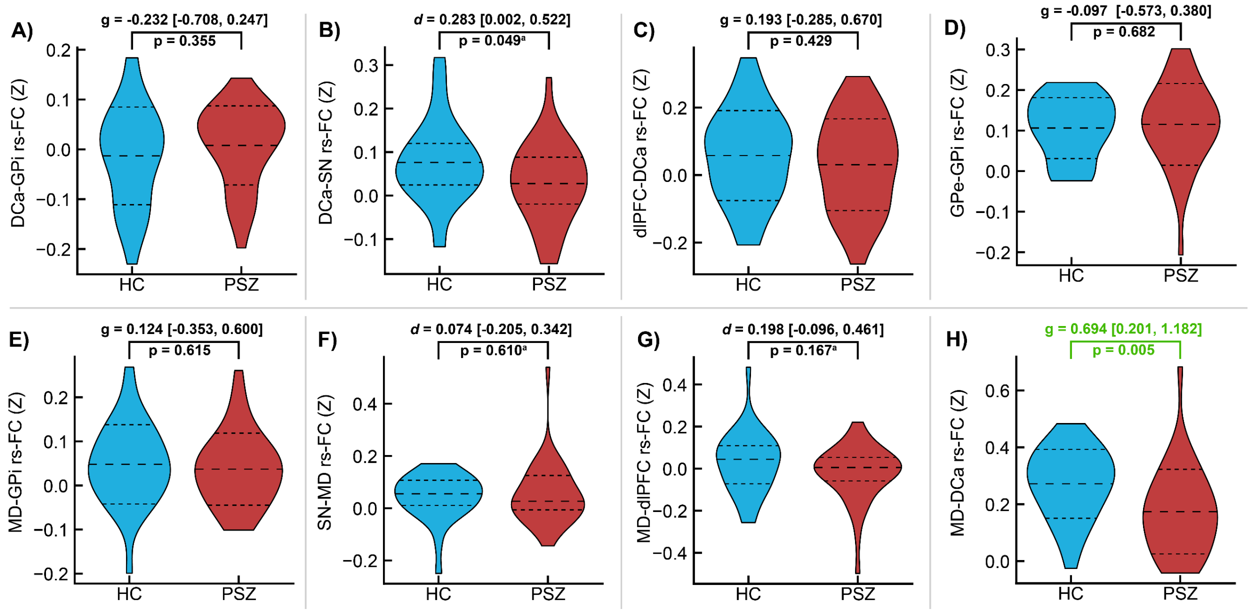


**Figure S6.** Group differences in pairwise, resting-state functional connectivity (rs-FC) between components of basal ganglia-thalamo-cortical circuitry between healthy controls (HC; n = 30; blue) and persons with schizophrenia (PSZ; n = 37; red). Connectivity in each pair was calculated while controlling for the average timeseries in all other network regions. Green text denotes statistical significance at p<0.05 after false discovery rate correction for multiple comparisons. Normally distributed data (as assessed using Lilliefors test) are shown with long-dashed and short-dashed lines denoting means and standard deviations, respectively, with statistical significance determined using independent samples t-tests and effect size calculated as Hedge's *g* **(Panels A, C-E, and H)**. Non-normally distributed data are shown with long-dashed and short-dashed lines denoting medians and quartiles, respectively, with statistical significance determined using Mann-Whitney U-tests (indicated by superscript letter a) and effect sizes calculated as Cliff's delta *d* **(Panels B, F, and G)**. DCa = dorsal caudate; GPi = globus pallidus internus; SN = substantia nigra; dlPFC = dorsolateral prefrontal cortex; GPe = globus pallidus externus; MD = mediodorsal nucleus; g = Hedges' *g* [95% Confidence Intervals]; *d* = Cliff's delta [95% Confidence Intervals].

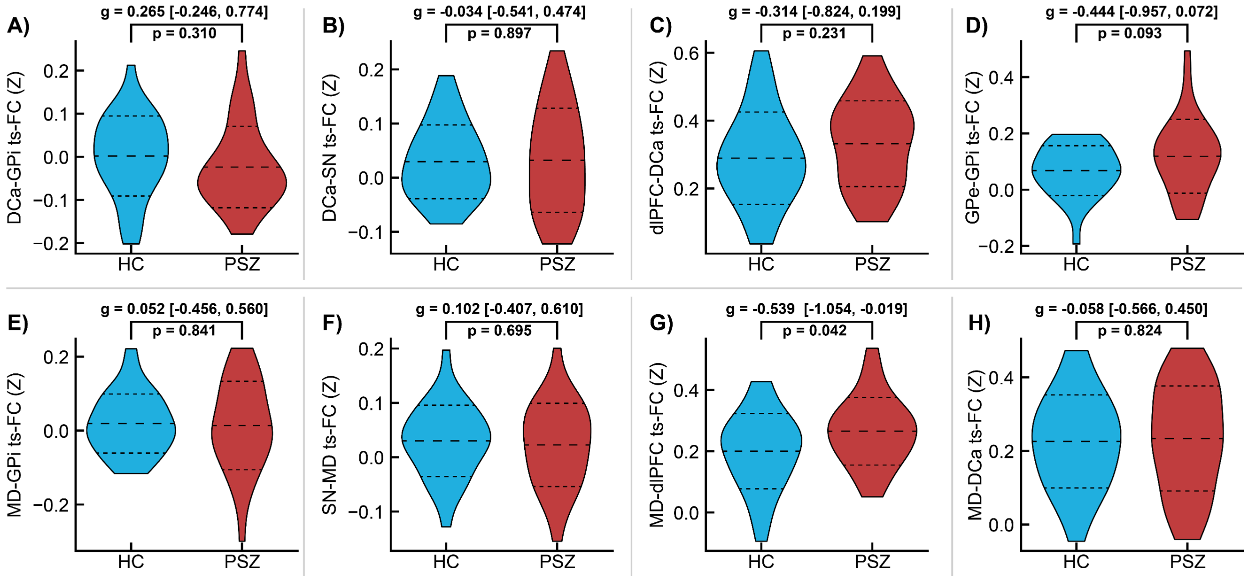


**Figure S7.** Group differences in pairwise, task-state functional connectivity (ts-FC) between components of basal ganglia-thalamo-cortical circuitry between healthy controls (HC; n = 29; blue) and persons with schizophrenia (PSZ; n = 29; red). Long-dashed and short-dashed lines denote means and standard deviations, respectively. Connectivity in each pair was calculated while controlling for the average timeseries in all other network regions. DCa = dorsal caudate; GPi = globus pallidus internus; SN = substantia nigra; dlPFC = dorsolateral prefrontal cortex; GPe = globus pallidus externus; MD = mediodorsal nucleus; g = Hedges' *g* [95% Confidence Intervals].

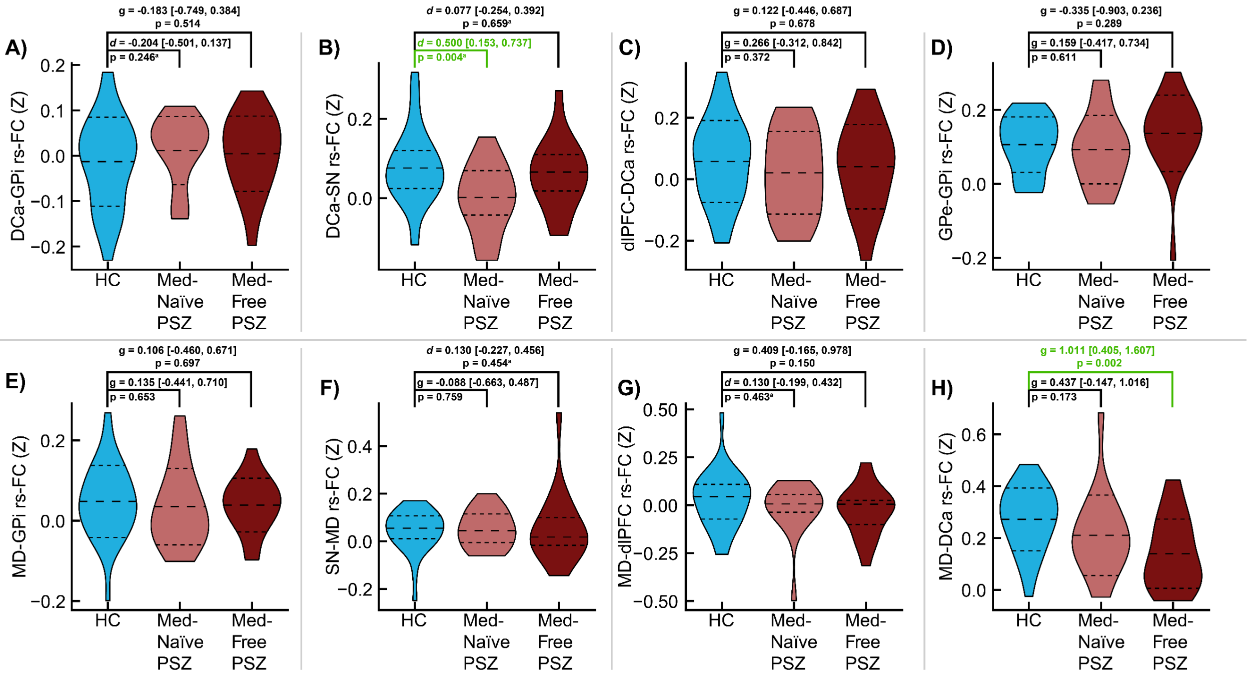


**Figure S8.** Differences in pairwise, resting-state functional connectivity (rs-FC) between components of basal ganglia-thalamo-cortical circuitry between healthy controls (n = 30) and medication-naïve (n = 15, light red) and medication-free (n = 14, dark red) persons with schizophrenia (PSZ), respectively. Connectivity in each pair was calculated while controlling for the average timeseries in all other network regions. Green text denotes statistical significance at p<0.05 after false discovery rate correction for multiple comparisons. Larger outer brackets denote comparisons between HC and medication-free PSZ, while smaller inner brackets denote comparisons between HC and medication-naïve PSZ. Normally distributed data (as assessed using Lilliefors test) are shown with long-dashed and short-dashed lines denoting means and standard deviations, respectively, with statistical significance determined using independent samples t-tests and effect size calculated as Hedge's *g*. Non-normally distributed data are shown with long-dashed and short-dashed lines denoting medians and quartiles, respectively, with statistical significance determined using Mann-Whitney U-tests (indicated by superscript letter a) and effect sizes calculated as Cliff's delta *d*. DCa = dorsal caudate; GPi = globus pallidus internus; SN = substantia nigra; dlPFC = dorsolateral prefrontal cortex; GPe = globus pallidus externus; MD = mediodorsal nucleus; g = Hedges' *g* [95% Confidence Intervals]; *d* = Cliff's delta [95% Confidence Intervals].

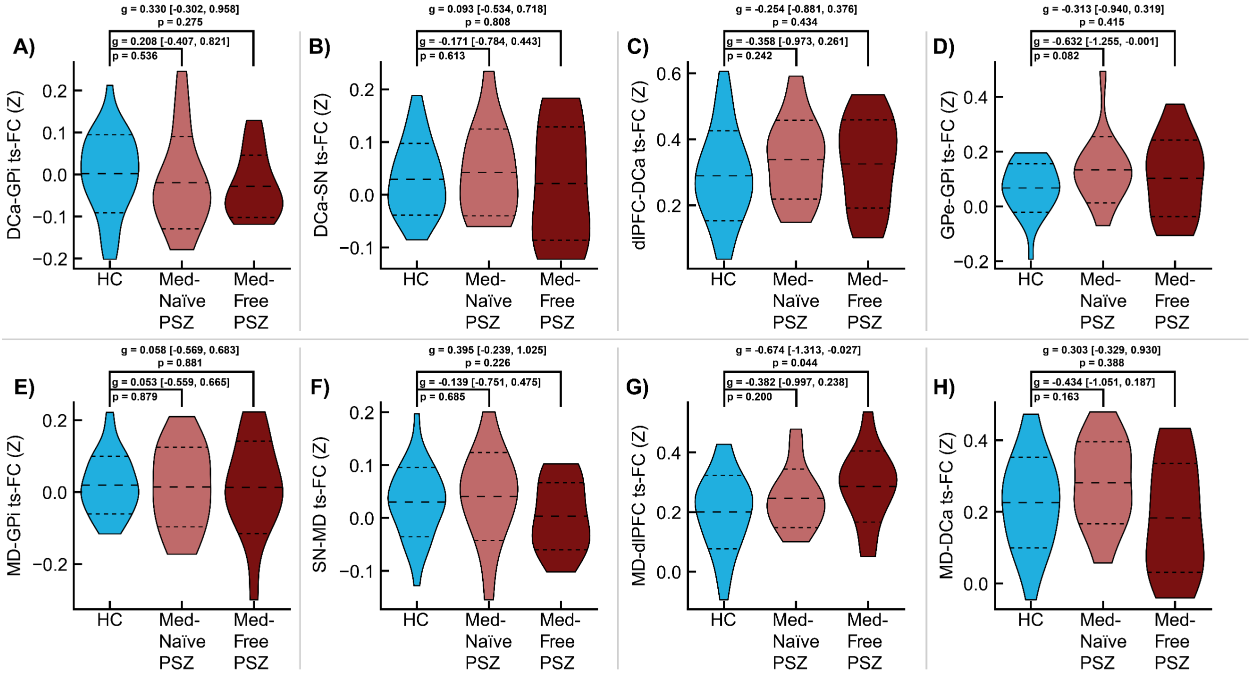


**Figure S9.** Differences in pairwise, task-state functional connectivity (ts-FC) between components of basal ganglia-thalamo-cortical circuitry between healthy controls (n = 30) and medication-naïve (n = 15, light red) and medication-free (n = 14, dark red) persons with schizophrenia (PSZ), respectively. Long-dashed and short-dashed lines denote means and standard deviations, respectively. Larger outer brackets denote comparisons between HC and medication-free PSZ, while smaller inner brackets denote comparisons between HC and medication-naïve PSZ. Connectivity in each pair was calculated while controlling for the average timeseries in all other network regions. DCa = dorsal caudate; GPi = globus pallidus internus; SN = substantia nigra; dlPFC = dorsolateral prefrontal cortex; GPe = globus pallidus externus; MD = mediodorsal nucleus; g = Hedges' *g* [95% Confidence Intervals].

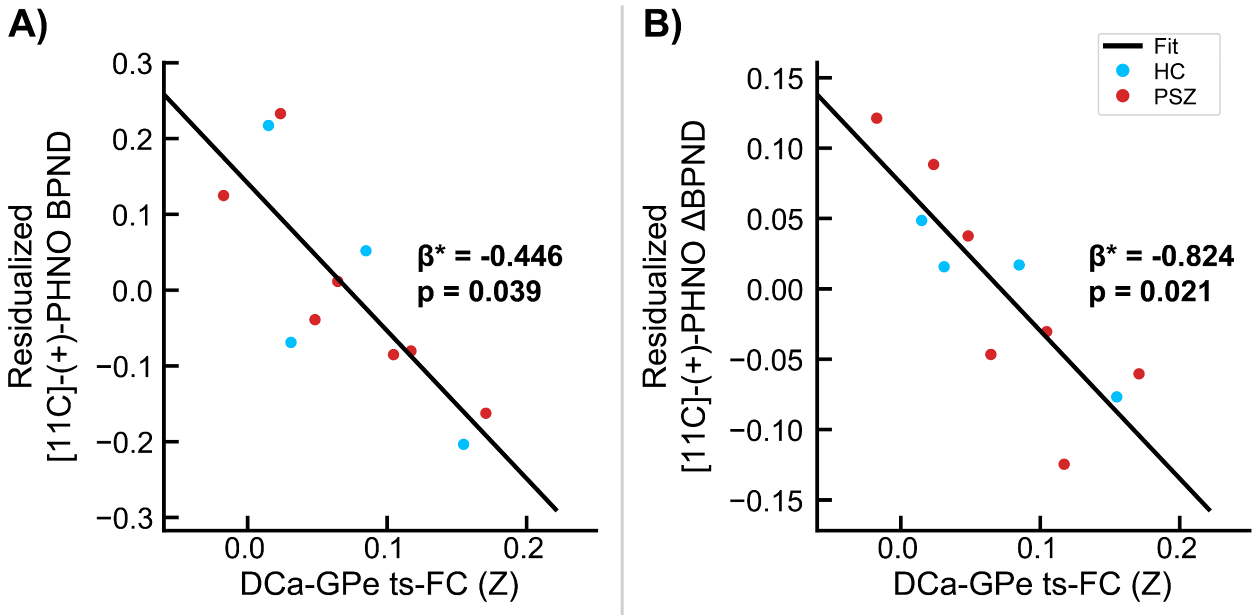


**Figure S10.** Associations between DCa-GPe ts-FC and **A)** [$^{11}$C]-(+)-PHNO Baseline BPND (N = 11) or **B)** ΔBPND (N = 11) amphetamine challenge, controlling for age, sex at birth, and diagnosis. Note that greater dopamine release is indicated by a more negative ΔBPND. Plot inlays show standardized regression coefficient β*. Blue dots denote healthy controls; red dots denote persons with schizophrenia.

### Table S1. Demographics for the Positron Emission Tomography Dataset

| Participant Characteristic | PSZ (n = 7) | HC (n = 4) | Effect Size [95% CIs] [a] | P-value |
|---|---|---|---|---|
| Age, Years (SD) | 23.1 (6.3) | 31.5 (11.9) | 0.89 [-0.33, 2.06][†] | 0.263[α] |
| Sex at Birth, Female (%) | 4 (57.1%) | 1 (25.0%) | 0.31 [0,0.95] | 0.303 |
| Ethnicity, Hispanic (%) | 0 (0.0%) | 1 (25.0%) | 0.42 [0, 1.05] | 0.165 |
| Race | | | 0.52 [0, 1.11 | 0.402 |
| African American (%) | 3 (42.9%) | 2 (50.0%) | | |
| Asian (%) | 2 (28.6%) | 0 (0.0%) | | |
| Caucasian (%) | 2 (28.6%) | 1 (25.0%) | | |
| More than one (%) | 0 (0.0%) | 0 (0.0%) | | |
| Other (%) | 0 (0.0%) | 1 (25.0%) | | |
| Handedness[b] | | | 0.42 [0, 1.05] | 0.165 |
| Ambidextrous (%) | 0 (0.0%) | 0 (0.0%) | | |
| Left (%) | 0 (0.0%) | 1 (25.0%) | | |
| Right (%) | 7 (100.0%) | 3 (75.0%) | | |
| Smoker[c] (%) | 2 (28.6%) | 0 (0.0%) | -0.36 [-0.99, 0] | 0.237 |
| Parental SES[d] (SD) | 46.8 (15.8) | 34.6 (5.9) | -0.43 [-0.85, 0.32] | 0.315 |
| PANSS Total (SD) | 78.6 (21.5) | 33.5 (4.5) | -2.32 [-3.82, -0.74] [†] | **0.001**[α] |
| PANSS General (SD) | 39.7 (14.3) | 17.8 (2.9) | -1.70 [-3.03, -0.30] [†] | **0.006**[α] |
| PANSS Negative (SD) | 19.1 (3.8) | 8.8 (1.7) | -2.91 [-4.61, -1.15] [†] | **$1.72 \times 10^{-4}$** [α] |
| PANSS Positive (SD) | 19.7 (5.6) | 7.0 (0) | -2.52 [-4.09, -0.88] [†] | **0.001**[α] |
| CDRS (SD) | 6.4 (5.3) | 0.8 (1.5) | -0.71 [-0.94, -0.03] | 0.079 |
| AP Medication-Naïve[e] (%) | 7 (100%) | — | — | — |

[a]Effect sizes for continuous variables were calculated with Hedges's g or Cliff's delta *d,* dependent on data normality (determined via Lilliefors test with a null hypothesis that data originates from a normal distribution at significance level p<0.05). Effect sizes for dichotomous and categorical variables were calculated with phi coefficients and Cramér's V, respectively.
[b]Handedness was reported using the Edinburgh Handedness Inventory.
[c]Smoking status defined as current daily nicotine use.
[d]Mean Parental SES was reported using the Hollingshead Four Factor Index Scale of Socioeconomic Status scale.
[e]All PSZ in this study were antipsychotic-free, defined as no exposure to oral antipsychotic medication for at least 3 weeks, and no exposure to intramuscular antipsychotic medications for at least 6 months, prior to study participation. PSZ were considered to be medication-naïve if they had fewer than 2 weeks of cumulative lifetime exposure to antipsychotic medications, with no prior use of long-acting injectable antipsychotics.
[α] Indicates use of independent samples t-test, rather than Mann-Whitney U-test, to obtain p-value.
† Indicates effect sizes for continuous variable calculated as Hedges's g, rather than Cliff's delta *d*.
**Bold p-values** indicate significance at p<0.05 (uncorrected).

*Abbreviations:* PSZ: Persons with Schizophrenia; HC: Healthy Controls; SD: standard deviation; SES: socioeconomic status; PANSS: Positive and Negative Syndrome Scale; CDRS: Calgary Depression Scale for Schizophrenia; AP: Antipsychotic; CIs: Confidence Intervals.

**Table S2.** Regression model assessing group differences in DCa-GPe rs-FC with age and sex at birth as covariates.

| Effect | Estimate (*B*) | *SE* (*B*) | β* | $t_{63}$ | p |
|---|---|---|---|---|---|
| Intercept | 0.065 | 0.012 | – | -0.031 | 0.976 |
| Age (Years) | -0.024 | 0.012 | -0.201 | -1.932 | 0.058 |
| Sex at Birth (Male) | 0.016 | 0.012 | 0.133 | 1.278 | 0.206 |
| Diagnosis (Schizophrenia) | 0.014 | 0.012 | 0.118 | 1.152 | 0.254 |

*Note.* N = 67, Error degrees of freedom = 63, R-squared = 0.085, Adjusted R-squared = 0.041, F-statistic versus constant model =1.94, p-value = 0.132. Model coefficients were estimated using robust regression. *Abbreviations*: ts-FC: task-state functional connectivity; *B*: unstandardized model coefficient; SE: standard error of unstandardized coefficient; β*: standardized model coefficient; N: number of observations; DCa: dorsal caudate; GPe: globus pallidus externus.

**Table S3.** Regression model assessing group differences in DCa-GPe ts-FC with age and sex at birth as covariates.

| Effect | Estimate (*B*) | *SE* (*B*) | β* | $t_{53}$ | p |
|---|---|---|---|---|---|
| Intercept | 0.079 | 0.013 | – | -0.253 | 0.802 |
| Age (Years) | 0.017 | 0.014 | 0.167 | 1.202 | 0.235 |
| Sex at Birth (Male) | -0.023 | 0.014 | -0.226 | -1.626 | 0.110 |
| Diagnosis (Schizophrenia) | 0.027 | 0.014 | 0.275 | 2.023 | 0.048 |

*Note.* N = 58, Error degrees of freedom = 53, R-squared = 0.126, Adjusted R-squared = 0.078, F-statistic versus constant model =2.600, p-value = 0.062. Model coefficients were estimated using robust regression.
*Abbreviations*: ts-FC: task-state functional connectivity; *B*: unstandardized model coefficient; SE: standard error of unstandardized coefficient; β*: standardized model coefficient; N: number of observations; DCa: dorsal caudate; GPe: globus pallidus externus.

**Table S4.** Regression model assessing group differences in DCa-GPe ts-FC with age, sex at birth, and smoking status as covariates.

| Effect | Estimate (*B*) | *SE* (*B*) | β* | $t_{52}$ | p |
|---|---|---|---|---|---|
| Intercept | 0.079 | 0.012 | – | -0.190 | 0.850 |
| Diagnosis (Schizophrenia) | 0.036 | 0.013 | 0.356 | 2.754 | 0.008 |
| Age (Years) | 0.017 | 0.013 | 0.166 | 1.300 | 0.199 |
| Sex at Birth (Male) | -0.019 | 0.013 | -0.185 | -1.410 | 0.165 |
| Smoking Status | -0.026 | 0.013 | -0.256 | -1.914 | 0.061 |

*Note.* N = 57, Error degrees of freedom = 52, R-squared = 0.212, Adjusted R-squared = 0.094, F-statistic versus constant model =3.51, p-value = 0.013. Model coefficients were estimated using robust regression.
*Abbreviations*: ts-FC: task-state functional connectivity; *B*: unstandardized model coefficient; SE: standard error of unstandardized coefficient; β*: standardized model coefficient; N: number of observations; DCa: dorsal caudate; GPe: globus pallidus externus.

**Table S5.** Regression model assessing group differences in DCa-GPe rs-FC with age, sex at birth, and smoking status as covariates.

| Effect | Estimate (*B*) | *SE* (*B*) | β* | $t_{60}$ | p |
|---|---|---|---|---|---|
| Intercept | 0.066 | 0.013 | – | 0.041 | 0.968 |
| Diagnosis (Schizophrenia) | 0.011 | 0.013 | 0.094 | 0.872 | 0.386 |
| Age (Years) | -0.022 | 0.013 | -0.181 | -1.701 | 0.094 |
| Sex at Birth (Male) | 0.014 | 0.013 | 0.117 | 1.081 | 0.284 |
| Smoking Status | 0.011 | 0.013 | 0.094 | 0.851 | 0.398 |

*Note.* N = 65, Error degrees of freedom = 60, R-squared = 0.083, Adjusted R-squared = 0.101, F-statistic versus constant model =1.35, p-value = 0.262. Model coefficients were estimated using robust regression.
*Abbreviations*: ts-FC: task-state functional connectivity; *B*: unstandardized model coefficient; SE: standard error of unstandardized coefficient; β*: standardized model coefficient; N: number of observations; DCa: dorsal caudate; GPe: globus pallidus externus.

**Table S6.** Regression model predicting average neuromelanin contrast-to-noise ratio in voxels within the substantia nigra previously found to be associated with psychosis severity, from the following predictors: intercept, age, sex at birth, diagnosis, and DCa-GPe ts-FC.

| Effect | Estimate (*B*) | *SE* (*B*) | β* | $t_{37}$ | p |
|---|---|---|---|---|---|
| Intercept | 8.691 | 1.371 | – | -0.475 | 0.638 |
| Age (Years) | 0.002 | 0.041 | 0.007 | 0.046 | 0.963 |
| Sex at Birth (Male) | -0.629 | 1.006 | -0.100 | -0.625 | 0.536 |
| Diagnosis (Schizophrenia) | -0.476 | 0.988 | -0.079 | -0.481 | 0.633 |
| DCa-GPe ts-FC | 11.272 | 4.748 | 0.395 | 2.376 | 0.023 |

*Note.* N = 42, Error degrees of freedom = 37, R-squared = 0.174, Adjusted R-squared = 0.085, F-statistic versus constant model = 1.950, p-value = 0.123. Model coefficients were estimated using robust regression. *Abbreviations*: ts-FC: task-state functional connectivity; *B*: unstandardized model coefficient; SE: standard error of unstandardized coefficient; β*: standardized model coefficient; N: number of observations; DCa: dorsal caudate; GPe: globus pallidus externus.

**Table S7.** Regression model predicting average neuromelanin contrast-to-noise ratio in voxels within the substantia nigra previously found to be associated with psychosis severity, from the following predictors: intercept, age, sex at birth, diagnosis, smoking status, and DCa-GPe ts-FC.

| Effect | Estimate (*B*) | *SE* (*B*) | β* | $t_{36}$ | p |
|---|---|---|---|---|---|
| Intercept | 9.058 | 0.455 | – | -0.361 | 0.720 |
| Diagnosis (Schizophrenia) | -0.295 | 0.528 | -0.097 | -0.558 | 0.580 |
| Age (Years) | 0.034 | 0.500 | 0.011 | 0.068 | 0.946 |
| Sex at Birth (Male) | -0.369 | 0.504 | -0.121 | -0.732 | 0.469 |
| Smoking Status | 0.111 | 0.518 | 0.036 | 0.214 | 0.832 |
| DCa-GPe ts-FC | 1.240 | 0.552 | 0.406 | 2.246 | 0.031 |

*Note.* N = 42, Error degrees of freedom = 36, R-squared = 0.175, Adjusted R-squared = 0.060, F-statistic versus constant model = 1.53, p-value = 0.206. Model coefficients were estimated using robust regression.
*Abbreviations*: ts-FC: task-state functional connectivity; *B*: unstandardized model coefficient; SE: standard error of unstandardized coefficient; β*: standardized model coefficient; N: number of observations; DCa: dorsal caudate; GPe: globus pallidus externus.

**Table S8.** Regression models in each group separately predicting average neuromelanin contrast-to-noise ratio in voxels within the substantia nigra previously found to be associated with psychosis severity, from the following predictors: intercept, age, sex at birth, and DCa-GPe ts-FC.

| **Healthy Participants**[a] | | | | | |
|---|---|---|---|---|---|
| **Effect** | **Estimate (*B*)** | ***SE* (*B*)** | **β*** | **$t_{20}$** | **p** |
| Intercept | 8.977 | 0.599 | – | -0.171 | 0.866 |
| Age (Years) | 1.152 | 0.637 | 0.398 | 1.810 | 0.089 |
| Sex at Birth (Male) | -1.088 | 0.645 | -0.376 | -1.687 | 0.111 |
| DCa-GPe ts-FC | 0.606 | 0.629 | 0.209 | 0.963 | 0.350 |
| **People with Schizophrenia**[b] | | | | | |
| **Effect** | **Estimate (*B*)** | ***SE* (*B*)** | **β*** | **$t_{22}$** | **p** |
| Intercept | 9.176 | 0.640 | – | -0.276 | 0.786 |
| Age (Years) | -0.701 | 0.708 | -0.216 | -0.991 | 0.335 |
| Sex at Birth (Male) | 0.430 | 0.728 | 0.132 | 0.591 | 0.562 |
| DCa-GPe ts-FC | 1.805 | 0.700 | 0.555 | 2.280 | 0.019 |

[a]N = 20, Error degrees of freedom = 16, R-squared = 0.280, Adjusted R-squared = 0.145, F-statistic versus constant model = 2.08, p-value = 0.143.

[b]N = 22, Error degrees of freedom = 18, R-squared = 0.281, Adjusted R-squared = 0.162, F-statistic versus constant model = 2.35, p-value = 0.107.

Model coefficients were estimated using robust regression.

*Abbreviations*: ts-FC: task-state functional connectivity; *B*: unstandardized model coefficient; SE: standard error of unstandardized coefficient; β*: standardized model coefficient; N: number of observations; DCa: dorsal caudate; GPe: globus pallidus externus.

**Table S9.** Regression models in each group separately predicting average neuromelanin contrast-to-noise ratio in voxels within the substantia nigra previously found to be associated with psychosis severity, from the following predictors: intercept, age, sex at birth, diagnosis, smoking status, and DCa-GPe ts-FC.

| | | | | | |
|---|---|---|---|---|---|
| **Healthy Participants[a]** | | | | | |
| **Effect** | **Estimate (*B*)** | ***SE* (*B*)** | **β*** | **$t_{20}$** | **p** |
| Intercept | 9.037 | 0.630 | – | -0.068 | 0.947 |
| Age (Years) | 1.270 | 0.669 | 0.439 | 1.898 | 0.077 |
| Sex at Birth (Male) | -1.176 | 0.679 | -0.406 | -1.732 | 0.104 |
| Smoking Status | -0.699 | 0.696 | -0.242 | -1.004 | 0.331 |
| DCa-GPe ts-FC | 0.334 | 0.711 | 0.115 | 0.470 | 0.645 |
| **People with Schizophrenia[b]** | | | | | |
| **Effect** | **Estimate (*B*)** | ***SE* (*B*)** | **β*** | **$t_{22}$** | **p** |
| Intercept | 9.141 | 0.621 | – | -0.340 | 0.768 |
| Age (Years) | -0.785 | 0.689 | -0.241 | -1.140 | 0.270 |
| Sex at Birth (Male) | 0.159 | 0.757 | 0.049 | 0.210 | 0.837 |
| Smoking Status | 0.634 | 0.761 | 0.195 | 0.833 | 0.416 |
| DCa-GPe ts-FC | 2.067 | 0.720 | 0.635 | 2.871 | 0.011 |

[a]N = 20, Error degrees of freedom = 15, R-squared = 0.323, Adjusted R-squared = 0.143, F-statistic versus constant model = 1.79, p-value = 0.183.

[b]N = 22, Error degrees of freedom = 17, R-squared = 0.343, Adjusted R-squared = 0.188, F-statistic versus constant model = 2.21, p-value = 0.111.

Model coefficients were estimated using robust regression.

*Abbreviations*: ts-FC: task-state functional connectivity; *B*: unstandardized model coefficient; SE: standard error of unstandardized coefficient; β*: standardized model coefficient; N: number of observations; DCa: dorsal caudate; GPe: globus pallidus externus.

**Table S10.** Regression model predicting average neuromelanin contrast-to-noise ratio in voxels within the substantia nigra previously found to be associated with psychosis severity, from the following predictors: intercept, age, sex at birth, diagnosis, DCa-GPe ts-FC and the interaction between diagnosis and DCa-GPe ts-FC.

| Effect | Estimate (*B*) | *SE* (*B*) | β* | $t_{52}$ | p |
|---|---|---|---|---|---|
| Intercept | 8.866 | 0.473 | – | -0.523 | 0.604 |
| Age (Years) | -0.056 | 0.480 | -0.018 | -0.116 | 0.908 |
| Sex at Birth (Male) | -0.285 | 0.475 | -0.093 | -0.600 | 0.552 |
| Diagnosis (Schizophrenia) | -0.168 | 0.491 | -0.055 | -0.342 | 0.734 |
| DCa-GPe ts-FC | 1.175 | 0.504 | 0.385 | 2.333 | 0.025 |
| Diagnosis Status * DCa-GPe ts-FC | 0.347 | 0.503 | 0.103 | 0.691 | 0.494 |

*Note.* N = 42, Error degrees of freedom = 36, R-squared = 0.20, Adjusted R-squared = 0.09, F-statistic versus constant model = 1.85, p-value = 0.129. Model coefficients were estimated using robust regression.
*Abbreviations*: ts-FC: task-state functional connectivity; *B*: unstandardized model coefficient; SE: standard error of unstandardized coefficient; β*: standardized model coefficient; N: number of observations; DCa: dorsal caudate; GPe: globus pallidus externus.

**Table S11.** Regression model in people with schizophrenia predicting average neuromelanin contrast-to-noise ratio in voxels within the substantia nigra previously found to be associated with psychosis severity from the following predictors: intercept, age, sex at birth, medication exposure (antipsychotic-free or antipsychotic-naïve), DCa-GPe ts-FC, and the interaction between medication status and ts-FC.

| **Effect** | **Estimate (*B*)** | ***SE* (*B*)** | **β*** | **$t_{16}$** | **p** |
|---|---|---|---|---|---|
| Intercept | 9.206 | 0.646 | – | -0.227 | 0.823 |
| Age (Years) | -0.824 | 0.975 | -0.253 | -0.845 | 0.411 |
| Sex at Birth (Male) | 0.278 | 0.785 | 0.085 | 0.354 | 0.728 |
| Medication Exposure (Antipsychotic-Free) | 0.295 | 0.953 | 0.091 | 0.309 | 0.761 |
| DCa-GPe ts-FC | 1.712 | 0.755 | 0.526 | 2.268 | 0.038 |
| Medication Status * DCa-GPe ts-FC | 0.140 | 0.751 | 0.043 | 0.187 | 0.854 |

*Note.* N = 22, Error degrees of freedom = 16, R-squared = 0.30, Adjusted R-squared = 0.081, F-statistic versus constant model = 1.37, p-value = 0.287. Model coefficients were estimated using robust regression. *Abbreviations*: SOT *k*: self-ordered working memory task working memory capacity; ts-FC: task-state functional connectivity; *B*: unstandardized model coefficient; SE: standard error of unstandardized coefficient; β*: standardized model coefficient; N: number of observations; DCa: dorsal caudate; GPe: globus pallidus externus.

**Table S12.** Mixed repeated-measures analysis of covariance with between-subject factors of diagnosis and average neuromelanin contrast-to-noise ratio in psychosis-associated substantia nigra voxels, a within-subject factor of DCa-GPe rs-FC or ts-FC (denoted as Condition), and age and sex at birth as covariates.

| **Effect** | **F(1, 36)** | **p** | **$\eta_p^2$** |
|---|---|---|---|
| Diagnosis (Schizophrenia) | 11.939 | 0.001 | 0.249 |
| Sex at Birth (Male) | 0.280 | 0.600 | 0.008 |
| Age | 2.723 | 0.108 | 0.070 |
| NM CNR | 0.002 | 0.961 | 0.00007 |
| NM CNR * Diagnosis | 0.200 | 0.658 | 0.006 |
| Diagnosis * Condition | 0.007 | 0.934 | 0.0002 |
| Sex at Birth * Condition | 0.337 | 0.565 | 0.009 |
| Age * Condition | 0.904 | 0.348 | 0.025 |
| NM CNR * Condition | 6.493 | 0.015 | 0.153 |
| NM CNR * Diagnosis * Condition | 2.388 | 0.131 | 0.062 |

*Note.* N = 42, Error degrees of freedom = 36. All effects tested with F(1, 36). Partial $\eta^2$ calculated as $SS_{effect}$ / ($SS_{effect}$ + $SS_{error}$).
Abbreviations: rs-FC: resting-state functional connectivity; ts-FC: task-state functional connectivity; DCa: dorsal caudate; GPe: globus pallidus externus; NM CNR: neuromelanin contrast-to-noise ratio.

**Table S13.** Regression models predicting [$^{11}$C]-(+)-PHNO baseline BPND and ΔBPND from the following predictors: Intercept, Age, Sex at Birth, Diagnosis, and DCa-GPe ts-FC.

| **Y = [$^{11}$C]-(+)-PHNO Baseline BPND[a]** | | | | | |
|---|---|---|---|---|---|
| **Effect** | **Estimate (*B*)** | ***SE* (*B*)** | **β*** | **$t_{11}$** | **p** |
| Intercept | 2.253 | 0.036 | – | 0.003 | 0.997 |
| Diagnosis (Schizophrenia) | 0.041 | 0.045 | 0.158 | 0.911 | 0.397 |
| Age (Years) | -0.253 | 0.050 | -0.970 | -5.057 | 0.002 |
| Sex at Birth (Male) | 0.011 | 0.041 | 0.040 | 0.261 | 0.803 |
| DCa-GPe ts-FC | -0.117 | 0.044 | -0.446 | -2.629 | 0.039 |
| **Y = [$^{11}$C]-(+)-PHNO ΔBPND[b]** | | | | | |
| **Effect** | **Estimate (*B*)** | ***SE* (*B*)** | **β*** | **$t_{11}$** | **p** |
| Intercept | -0.125 | 0.017 | – | 0.056 | 0.957 |
| Diagnosis (Schizophrenia) | -0.056 | 0.021 | -0.734 | -2.695 | 0.036 |
| Age (Years) | -0.059 | 0.023 | -0.772 | -2.569 | 0.042 |
| Sex at Birth (Male) | 0.002 | 0.019 | 0.027 | 0.113 | 0.914 |
| DCa-GPe ts-FC | -0.063 | 0.020 | -0.824 | -3.098 | 0.021 |

[a]N = 11, Error degrees of freedom = 6, R-squared = 0.874, Adjusted R-squared = 0.790, F-statistic versus constant model = 10.400, p-value = 0.007.

[b]N = 11, Error degrees of freedom = 6, R-squared = 0.701, Adjusted R-squared = 0.501, F-statistic versus constant model = 3.510, p-value = 0.083.

Model coefficients were estimated using robust regression.

*Abbreviations*: ts-FC: task-state functional connectivity; *B*: unstandardized model coefficient; SE: standard error of unstandardized coefficient; β*: standardized model coefficient; N: number of observations; DCa: dorsal caudate; GPe: globus pallidus externus; BPND: Non-displaceable binding potential.

**Table S14.** Regression models predicting [$^{11}$C]-(+)-PHNO baseline BPND and ΔBPND from the following predictors: Intercept, Age, Sex at Birth, Diagnosis, Smoking Status and DCa-GPe ts-FC.

| **Y = [$^{11}$C]-(+)-PHNO Baseline BPND[a]** | | | | | |
|---|---|---|---|---|---|
| **Effect** | **Estimate (*B*)** | ***SE* (*B*)** | **β*** | **$t_5$** | **p** |
| Intercept | 2.253 | 0.064 | – | -0.004 | 0.997 |
| Diagnosis (Schizophrenia) | 0.152 | 0.112 | 0.582 | 1.136 | 0.233 |
| Age (Years) | -0.160 | 0.080 | -0.614 | -2.000 | 0.102 |
| Sex at Birth (Male) | 0.019 | 0.096 | 0.073 | 0.199 | 0.850 |
| Smoking Status | -0.038 | 0.095 | -0.147 | -0.403 | 0.704 |
| DCa-GPe ts-FC | -0.076 | 0.099 | -0.292 | -0.769 | 0.476 |
| **Y = [$^{11}$C]-(+)-PHNO ΔBPND[b]** | | | | | |
| **Effect** | **Estimate (*B*)** | ***SE* (*B*)** | **β*** | **$t_{11}$** | **p** |
| Intercept | -0.126 | 0.026 | – | $-1.52 \times 10^{-16}$ | 0.999 |
| Diagnosis (Schizophrenia) | -0.0004 | 0.045 | -0.006 | -0.009 | 0.993 |
| Age (Years) | -0.008 | 0.032 | -0.110 | -0.259 | 0.806 |
| Sex at Birth (Male) | 0.004 | 0.039 | 0.050 | 0.099 | 0.925 |
| Smoking Status | -0.023 | 0.038 | -0.302 | -0.601 | 0.574 |
| DCa-GPe ts-FC | -0.037 | 0.040 | -0.491 | -0.936 | 0.392 |

[a]N = 11, Error degrees of freedom = 5, R-squared = 0.699, Adjusted R-squared = 0.398, F-statistic versus constant model = 2.32, p-value = 0.188.

[b]N = 11, Error degrees of freedom = 5, R-squared = 0.380, Adjusted R-squared = -0.240, F-statistic versus constant model = 0.613, p-value = 0.698.

Model coefficients were estimated using robust regression.

*Abbreviations*: ts-FC: task-state functional connectivity; *B*: unstandardized model coefficient; SE: standard error of unstandardized coefficient; β*: standardized model coefficient; N: number of observations; DCa: dorsal caudate; GPe: globus pallidus externus; BPND: Non-displaceable binding potential.

**Table S15.** Regression model predicting SOT *k* from the following predictors: intercept, age, sex at birth, diagnosis, and DCa-GPe ts-FC.

| Effect | Estimate (*B*) | *SE* (*B*) | β* | $t_{53}$ | p |
|---|---|---|---|---|---|
| Intercept | 5.018 | 0.244 | – | 0.252 | 0.802 |
| Age (Years) | -0.791 | 0.258 | -0.380 | -3.061 | 0.004 |
| Sex at Birth (Male) | 0.267 | 0.262 | 0.128 | 1.019 | 0.313 |
| Diagnosis (Schizophrenia) | -0.352 | 0.259 | -0.169 | -1.360 | 0.180 |
| DCa-GPe ts-FC | -0.651 | 0.271 | -0.313 | -2.405 | 0.020 |

*Note.* N = 58, Error degrees of freedom = 53, R-squared = 0.326, Adjusted R-squared = 0.275, F-statistic versus constant model = 6.396, p-value = $2.819 \times 10^{-4}$. Model coefficients were estimated using robust regression. *Abbreviations*: SOT *k*: self-ordered working memory task working memory capacity; ts-FC: task-state functional connectivity; *B*: unstandardized model coefficient; SE: standard error of unstandardized coefficient; β*: standardized model coefficient; N: number of observations; DCa: dorsal caudate; GPe: globus pallidus externus.

**Table S16.** Regression model predicting SOT *k* from the following predictors: intercept, age, sex at birth, diagnosis, smoking status, and DCa-GPe ts-FC.

| Effect | Estimate (*B*) | *SE* (*B*) | β* | $t_{51}$ | p |
|---|---|---|---|---|---|
| Intercept | 4.999 | 0.241 | – | 0.239 | 0.812 |
| Diagnosis (Schizophrenia) | -0.225 | 0.271 | -0.107 | -0.829 | 0.411 |
| Age (Years) | -0.814 | 0.256 | -0.389 | -3.185 | 0.002 |
| Sex at Birth (Male) | 0.301 | 0.262 | 0.144 | 1.147 | 0.257 |
| Smoking Status | -0.344 | 0.272 | -0.164 | -1.265 | 0.212 |
| DCa-GPe ts-FC | -0.770 | 0.279 | -0.368 | -2.762 | 0.008 |

*Note.* N = 57, Error degrees of freedom = 51, R-squared = 0.372, Adjusted R-squared = 0.311, F-statistic versus constant model = 6.05, p-value = 0.0002. Model coefficients were estimated using robust regression. *Abbreviations*: SOT *k*: self-ordered working memory task working memory capacity; ts-FC: task-state functional connectivity; *B*: unstandardized model coefficient; SE: standard error of unstandardized coefficient; β*: standardized model coefficient; N: number of observations; DCa: dorsal caudate; GPe: globus pallidus externus.

**Table S17.** Regression models predicting SOT *k* separately in each group from the following predictors: Intercept, Age, Sex at Birth, and DCa-GPe ts-FC.

| **Healthy Participants[a]** | | | | | |
|---|---|---|---|---|---|
| **Effect** | **Estimate (*B*)** | ***SE* (*B*)** | **β*** | **$t_{29}$** | **p** |
| Intercept | 5.611 | 0.295 | – | 0.333 | 0.742 |
| Age (Years) | -0.029 | 0.314 | -0.018 | -0.094 | 0.926 |
| Sex at Birth (Male) | 0.506 | 0.317 | 0.310 | 1.597 | 0.123 |
| DCa-GPe ts-FC | -0.484 | 0.313 | -0.297 | -1.548 | 0.134 |
| **People with Schizophrenia[b]** | | | | | |
| **Effect** | **Estimate (*B*)** | ***SE* (*B*)** | **β*** | **$t_{29}$** | **p** |
| Intercept | 4.366 | 0.333 | – | -0.105 | 0.917 |
| Age (Years) | -1.407 | 0.356 | -0.600 | -3.951 | 0.0006 |
| Sex at Birth (Male) | -0.052 | 0.364 | -0.022 | -0.144 | 0.887 |
| DCa-GPe ts-FC | -0.836 | 0.360 | -0.356 | -2.324 | 0.029 |

[a]N = 29, Error degrees of freedom = 25, R-squared = 0.208, Adjusted R-squared = 0.113, F-statistic versus constant model = 2.19, p-value = 0.114.

[b]N = 29, Error degrees of freedom = 25, R-squared = 0.516, Adjusted R-squared = 0.458, F-statistic versus constant model = 8.87, p-value = 0.0004.

Model coefficients were estimated using robust regression.

*Abbreviations*: ts-FC: task-state functional connectivity; *B*: unstandardized model coefficient; SE: standard error of unstandardized coefficient; β*: standardized model coefficient; N: number of observations; DCa: dorsal caudate; GPe: globus pallidus externus.

**Table S18.** Regression models predicting SOT *k* separately in each group from the following predictors: Intercept, Age, Sex at Birth, Smoking Status and DCa-GPe ts-FC.

| Effect | Estimate (*B*) | *SE* (*B*) | β* | $t_{29}$ | p |
|---|---|---|---|---|---|
| **Healthy Participants[a]** | | | | | |
| Intercept | 5.577 | 0.290 | – | 0.222 | 0.826 |
| Age (Years) | -0.019 | 0.308 | -0.011 | -0.060 | 0.952 |
| Sex at Birth (Male) | 0.532 | 0.312 | 0.326 | 1.707 | 0.101 |
| Smoking Status | 0.154 | 0.314 | 0.095 | 0.491 | 0.628 |
| DCa-GPe ts-FC | -0.430 | 0.326 | -0.263 | -1.318 | 0.200 |
| **People with Schizophrenia[b]** | | | | | |
| **Effect** | **Estimate (*B*)** | ***SE* (*B*)** | **β*** | **$t_{29}$** | **p** |
| Intercept | 4.377 | 0.346 | – | -0.069 | 0.945 |
| Age (Years) | -1.370 | 0.375 | -0.584 | -3.654 | 0.001 |
| Sex at Birth (Male) | 0.181 | 0.406 | 0.077 | 0.446 | 0.660 |
| Smoking Status | -0.539 | 0.409 | -0.230 | -1.318 | 0.200 |
| DCa-GPe ts-FC | -0.929 | 0.385 | -0.396 | -2.410 | 0.024 |

[a]N = 29, Error degrees of freedom = 24, R-squared = 0.231, Adjusted R-squared = 0.103, F-statistic versus constant model = 1.8, p-value = 0.162.

[b]N = 29, Error degrees of freedom = 24, R-squared = 0.528, Adjusted R-squared = 0.449, F-statistic versus constant model = 6.7, p-value = 0.0009.

Model coefficients were estimated using robust regression.

*Abbreviations*: ts-FC: task-state functional connectivity; *B*: unstandardized model coefficient; SE: standard error of unstandardized coefficient; β*: standardized model coefficient; N: number of observations; DCa: dorsal caudate; GPe: globus pallidus externus.

**Table S19.** Regression model predicting SOT *k* from the following predictors: intercept, age, sex at birth, diagnosis, DCa-GPe ts-FC and the interaction between diagnosis and DCa-GPe ts-FC.

| Effect | Estimate (*B*) | *SE* (*B*) | β* | $t_{52}$ | p |
|---|---|---|---|---|---|
| Intercept | 5.130 | 0.246 | – | 0.427 | 0.671 |
| Age (Years) | -0.716 | 0.248 | -0.344 | -2.882 | 0.006 |
| Sex at Birth (Male) | 0.248 | 0.252 | 0.119 | 0.986 | 0.329 |
| Diagnosis (Schizophrenia) | -0.348 | 0.249 | -0.167 | -0.140 | 0.168 |
| DCa-GPe ts-FC | -0.716 | 0.261 | -0.344 | -2.746 | 0.008 |
| Diagnosis Status * DCa-GPe ts-FC | -0.252 | 0.251 | -0.115 | -1.004 | 0.320 |

*Note.* N = 58, Error degrees of freedom = 52, R-squared = 0.36, Adjusted R-squared = 0.30, F-statistic versus constant model = 5.85, p-value = 0.0002. Model coefficients were estimated using robust regression. *Abbreviations*: SOT *k*: self-ordered working memory task working memory capacity; ts-FC: task-state functional connectivity; *B*: unstandardized model coefficient; SE: standard error of unstandardized coefficient; β*: standardized model coefficient; N: number of observations; DCa: dorsal caudate; GPe: globus pallidus externus.

**Table S20.** Regression model predicting SOT *k* in people with schizophrenia, only, from the following predictors: intercept, age, sex at birth, medication status (antipsychotic-free or antipsychotic-naïve), DCa-GPe ts-FC, and the interaction between medication exposure and ts-FC.

| Effect | Estimate (*B*) | *SE* (*B*) | β* | $t_{29}$ | p |
|---|---|---|---|---|---|
| Intercept | 4.386 | 0.374 | – | -0.040 | 0.969 |
| Age (Years) | -0.923 | 0.483 | -0.394 | -1.910 | 0.069 |
| Sex at Birth (Male) | -0.249 | 0.420 | -0.106 | -0.593 | 0.559 |
| Medication Exposure (Antipsychotic-Free) | -0.329 | 0.469 | -0.140 | -0.702 | 0.490 |
| DCa-GPe ts-FC | -1.171 | 0.418 | -0.499 | -2.800 | 0.010 |
| Medication Status * DCa-GPe ts-FC | 0.133 | 0.429 | 0.057 | 0.310 | 0.760 |

*Note.* N = 29, Error degrees of freedom = 23, R-squared = 0.429, Adjusted R-squared = 0.305, F-statistic versus constant model = 3.46, p-value = 0.018. Model coefficients were estimated using robust regression. *Abbreviations*: SOT *k*: self-ordered working memory task working memory capacity; ts-FC: task-state functional connectivity; *B*: unstandardized model coefficient; SE: standard error of unstandardized coefficient; β*: standardized model coefficient; N: number of observations; DCa: dorsal caudate; GPe: globus pallidus externus.

**Table S21.** Mixed repeated-measures analysis of covariance with between-subject factors of diagnosis and SOT capacity *k*, a within-subject factor of DCa-GPe rs-FC or ts-FC (denoted as Condition), and age and sex at birth as covariates.

| Effect | F(1, 50) | p | $\eta_p^2$ |
|---|---|---|---|
| Diagnosis (Schizophrenia) | 7.460 | 0.009 | 0.130 |
| Sex at Birth (Male) | 0.523 | 0.473 | 0.010 |
| Age | 0.637 | 0.428 | 0.013 |
| SOT *k* | 0.320 | 0.574 | 0.006 |
| SOT *k* * Diagnosis | 0.004 | 0.953 | 0.00007 |
| Diagnosis * Condition | 0.240 | 0.626 | 0.005 |
| Sex at Birth * Condition | 0.216 | 0.644 | 0.004 |
| Age * Condition | 0.133 | 0.717 | 0.003 |
| SOT *k* * Condition | 4.364 | 0.042 | 0.080 |
| SOT *k* * Diagnosis * Condition | 1.095 | 0.300 | 0.021 |

*Note.* N = 56, Error degrees of freedom = 50. All effects tested with F(1, 50). Partial $\eta^2$ calculated as $SS_{effect}$ / ($SS_{effect}$ + $SS_{error}$).
Abbreviations: rs-FC: resting-state functional connectivity; ts-FC: task-state functional connectivity; DCa: dorsal caudate; GPe: globus pallidus externus.

**Table S22.** Regression models predicting PANSS Positive and Negative symptom scores in persons with schizophrenia from the following predictors: Intercept, Age, Sex at Birth, and DCa-GPe ts-FC.

| **Y = PANSS Positive**[a] | | | | | |
|---|---|---|---|---|---|
| **Effect** | **Estimate (*B*)** | ***SE* (*B*)** | **β*** | **$t_{27}$** | **p** |
| Intercept | 16.871 | 3.148 | – | -0.506 | 0.617 |
| Age (Years) | 0.049 | 0.085 | 0.117 | 0.570 | 0.574 |
| Sex at Birth (Male) | -1.088 | 2.358 | -0.096 | -0.462 | 0.649 |
| DCa-GPe ts-FC | -21.205 | 11.087 | -0.393 | -1.913 | 0.068 |
| **Y = PANSS Negative**[b] | | | | | |
| **Effect** | **Estimate (*B*)** | ***SE* (*B*)** | **β*** | **$t_{27}$** | **p** |
| Intercept | 17.777 | 3.161 | – | -0.193 | 0.849 |
| Age (Years) | -0.061 | 0.086 | -0.162 | -0.709 | 0.486 |
| Sex at Birth (Male) | -0.662 | 2.367 | -0.065 | -0.280 | 0.782 |
| DCa-GPe ts-FC | -9.773 | 11.133 | -0.201 | -0.878 | 0.389 |

[a]N = 27, Error degrees of freedom = 23, R-squared = 0.146, Adjusted R-squared = 0.035, F-statistic versus constant model = 1.313, p-value = 0.294.

[b]N = 27, Error degrees of freedom = 23, R-squared = 0.074, Adjusted R-squared = -0.047, F-statistic versus constant model = 0.613, p-value = 0.614.

Model coefficients were estimated using robust regression.

*Abbreviations*: ts-FC: task-state functional connectivity; *B*: unstandardized model coefficient; SE: standard error of unstandardized coefficient; β*: standardized model coefficient; N: number of observations; DCa: dorsal caudate; GPe: globus pallidus externus; PANSS: Positive and Negative Syndrome Scale.
